# New-onset type 2 diabetes mellitus and obesity-related cancer risk: a matched cohort study (UK Biobank)

**DOI:** 10.64898/2026.08.25.26360729

**Authors:** Owen Tipping, Mengying Wang, Richard M. Martin, Matthew Sperrin, Andrew G. Renehan

## Abstract

**Background:** Observational research reports positive associations between type 2 diabetes mellitus (T2DM) and obesity-related cancers (ORCs), but causality remains unclear due to confounding (namely the shared risk factor of obesity, commonly approximated as body mass index, BMI), immortal time bias, and detection-time bias. Here, we aimed to use causal inference methods to minimise the above problems and estimate causal associations between new-onset T2DM and incident cancer.

**Methods:** We performed a cohort study within UK Biobank, comparing new-onset T2DM with unexposed individuals matched 1 to 3 on BMI, age, and sex using a sequential longitudinal approach. The primary outcomes were total incident cancer, divided into ORCs and non-obesity-related cancers (NORCs). The secondary outcomes were site-specific cancers. We developed Cox models to estimate time-split hazard ratios (tsHRs) and 95% confidence intervals (CIs) stratified by sex.

**Findings:** 23,771 participants with new-onset T2DM were matched with 71,170 unexposed participants. During a median follow-up of 5 years, there were 7694 (T2DM: 2432; unexposed: 5262) incident cancers. In men, there was evidence for an effect of T2DM on obesity-related cancer (tsHR 1.39, 95% CI 1.21-1.59), particularly on hepatocellular carcinoma (tsHR 3.97, 95% CI 2.38-6.65), pancreatic (tsHR 1.77, 95% CI 1.15-2.72) and kidney (tsHR 1.62, 95% CI 1.13-2.32) cancers. In women, there was evidence for an effect on obesity-related cancers (tsHR 1.33, 95% CI 1.16-1.52). Importantly, there were no associations with post-menopausal breast and endometrial cancers, two cancer types consistently associated with elevated BMI. There was no effect of new-onset T2DM on incidence of NORCs. There was evidence of detection-time bias, particularly in men.

**Interpretation:** This is the first large-scale study to demonstrate evidence of a BMI-independent associations between new-onset T2DM and incident cancer. In men, this was primarily driven by hepatocellular carcinoma, pancreatic cancer, and kidney cancer. In women, the underlying cancers driving this relationship were less clearly defined.

**Funding:** This study was funded by Cancer Research UK and administered through the Manchester Cancer Research Centre MB-PhD scheme (SEBCATP-2023/100010).

**Novelty and impact statement:** This study addresses key limitations of previous research examining T2DM and cancer risk. Unlike earlier studies that modelled T2DM as a fixed exposure and were therefore susceptible to immortal time bias, a novel longitudinal matching algorithm was implemented and treated T2DM as a time-varying exposure. In addition, extensive matching and adjustment for anthropometric measures enabled estimation of obesity-independent effects, while a time-split Cox modelling framework was used to assess and minimise detection bias and reverse causality.

**Research in context:** *Evidence before this study:* We performed a literature search relevant to the research question and identified two umbrella reviews and six meta-analyses/ systematic reviews. The studies included within the reviews were primarily based on observational evidence, except for 1 umbrella review and meta-analysis that incorporated Mendelian Randomisation (MR) evidence. The literature demonstrated an overall positive association between T2DM and cancer, however, there were discrepancies between observational and MR evidence. There were consistent associations with multiple site-specific cancers from observational evidence: pancreatic, hepatocellular, post-menopausal breast, endometrial, and colorectal, as well as inverse associations with prostate cancer. However, MR evidence illustrated a lack of consistency with observational evidence. MR studies from an umbrella review identified evidence for a causal association between T2DM and pancreatic cancer, but limited evidence for endometrial, breast, and colorectal cancer. Furthermore, it remains unclear whether the associations identified in previous observational research are causal, or due to confounding (e.g., mutual risk factor of obesity); immortal time bias (many studies combine prevalent and new-onset T2DM); time-detection bias (e.g., the co-diagnosis of two relatively common conditions at the same time); or reverse causality (liver/pancreatic cancer causing diabetes onset, but being diagnosed after the initial diabetes diagnosis).

*Added value of this study:* Our study addressed the major issue of detection-time bias/reverse causality in previous T2DM studies by time-splitting at 1 year. Similarly, with BMI as a well-established mutual risk factor for T2DM and cancer, we performed exact matching on BMI and further adjusted for BMI as a covariate in our Cox models. We also performed sensitivity analyses, replacing BMI with alternative anthropometric measurements including waist circumference (WC), and waist-to-hip ratio (WHR). In contrast to previous studies using a never/ever categorisation for T2DM, we treated T2DM as a time-varying exposure, by adopting a sequential longitudinal matching algorithm using the date T2DM diagnosis as each participant’s study entry point, which helped to alleviate the effects of immortal time bias and allowed unexposed participants to develop T2DM and be subsequently followed up as an exposed participant.

*Implications of all the available evidence:* After adjusting for detection-time bias, reverse causality, immortal time bias, and key confounders (BMI, smoking, deprivation, physical activity, age, sex, and alcohol consumption), our study highlighted evidence for associations between T2DM and multiple site-specific cancers, with the strongest evidence for pancreatic, liver and kidney cancers. This remained consistent with associations established in prior studies. However, we found minimal evidence for an association between T2DM, endometrial, and post-menopausal breast cancer, which has been previously reported. Overall, our study supports the notion that T2DM may be causally associated with a small number of ORCs independent of obesity. Finally, our findings highlight the significant effect of detection-time bias/reverse causality in T2DM studies, which we observed to be stronger in men, and we caution researchers to ensure they appropriately adjust for these key biases whilst trying to elucidate causal associations with T2DM as an exposure.

## Introduction

A potential link between type 2 diabetes mellitus (T2DM), a predominantly obesity-related metabolic condition, and cancer has been recognised for several decades. Two umbrella reviews (1, 2) and six systematic reviews (3–8) that investigated associations between T2DM and either total incident cancers or specific cancer incidences have reported positive associations with the following obesity-related cancers (ORCs): hepatocellular carcinoma and pancreatic, post-menopausal breast, endometrial, kidney and colorectal cancers. These reviews also consistently report that T2DM is inversely associated with prostate cancer, which is a non-obesity-related cancer (NORC). Based on estimated effect sizes from these reviews, models calculate that, for all incident cancers, 8.5% in men and 4.8% in women are attributable to the effects of diabetes (9). Furthermore, cancer is overtaking cardiovascular complications as the leading cause of mortality in several populations with diabetes (10).

However, whether these associations are causal has been questioned at a number of levels (11). First, obesity, commonly approximated by body mass index (BMI) is casually associated with increased risk at least 13 site-specific cancers, which overlap many of the cancers associated with T2DM i.e. confounding (12). Several proposed mechanisms, operating in obesity, may be at play in T2DM – for example, mitogenic effects of hyperinsulinemia, increased levels of IGF1, hyperglycaemia, chronic inflammation and perturbations in bioavailable oestrogen (13, 14). Second, Mendelian randomization studies generally contradict the findings of conventional epidemiological studies, elucidating evidence for a positive causal effect with pancreatic cancer only (1). Third, many longitudinal studies model T2DM exposure as a fixed time-independent covariate according to baseline status and include participants with prevalent diabetes, an approach that is subject to *immortal time bias* and fails to account for unexposed participants becoming exposed during follow-up (15–17). Including participants diagnosed with diabetes prior to study entry requires them to have survived and remained event-free until enrolment, creating a period of ‘immortal time’ in which they cannot experience the outcome, leading to bias and an underestimation of outcome risk (18). Fourth, previous observational studies have also shown evidence of *detection-time bias* shortly after diabetes diagnosis where increased medical monitoring inflates post-diagnosis cancer incidence due to earlier detection. Similarly, researchers have highlighted the issue of reverse causality whereby pancreatic and liver cancer causes diabetes onset, but the initial cancers are diagnosed later (19).

Our study addressed these uncertainties using casual inference approaches. Specifically, we (i) limited our population to new-onset T2DM to minimise immortal time bias; (ii) matched on BMI to reduce confounding; (iii) matched using a sequential longitudinal method, again to minimise immortal time bias; (iv) used time-split Cox models at 1 year to account for detection-time bias; and (v) finally, subdivided cancers into obesity-related cancers (ORCs) and non-obesity-related cancers (NORCs), again to test whether obesity might be an important confounder in the links between T2DM and cancer.

## Methods

### Population

We performed a matched cohort study within the UK Biobank, a large prospective British cohort that recruited approximately 500,000 men and women aged 40-69 across England, Wales, and Scotland between 2006 and 2010 (20). Participants attended assessment centres, filled out subsequent health-related questionnaires, and underwent anthropometric measurements. Longitudinal follow-up was achieved through linkage to national datasets including the death registry, cancer registry, primary care, as well as hospital inpatient and outpatient records. UK Biobank has ethical approval from the Northwest Multi-centre Research Ethics Committee (MREC) as a Research Tissue Bank (RTB), and separate ethical approval for this study was not required. Participants attended assessment centres, filled out subsequent health-related questionnaires, and underwent anthropometric measurements. Longitudinal follow-up was achieved through linkage to national datasets including the death registry, cancer registry, primary care, as well as hospital inpatient and outpatient records. UK Biobank has ethical approval from the Northwest Multi-centre Research Ethics Committee (MREC) as a Research Tissue Bank. A separate ethical approval was not required for this study.

### New-onset type 2 diabetes cohort

To minimise immortal time bias, we identified participants with new-onset T2DM, defined by the date of first reported non-insulin dependent diabetes mellitus (E11) data field. Dates were primarily sourced from linked hospital admissions data, followed by primary care records, self-reported data, and the death register. We excluded participants with prevalent diabetes date prior to UK Biobank enrolment; a date of diagnosis of diabetes after the end of study date (censoring date); a past medical history of cancer; missing baseline BMI measurements; or lost to follow-up due to withdrawing consent, leaving the country, or reported death without an official death certificate.

### Outcome measure

The primary outcome was total cancer incidence sub-classified into obesity-related (ORCs) and non-obesity-related cancers (NORCs). Cancer outcomes were defined by the first occurrence of a malignant neoplasm in the cancer registry (ICD-10 C00-C97) following UK Biobank recruitment. From this definition, we excluded non-melanotic skin cancer (C44), carcinoma in situ (D00–D09), benign neoplasms (D10–D36), benign neuroendocrine tumours (D3A), neoplasms of uncertain behaviour (D37–D48), as well as neoplasms of unspecified behaviour (D49), with the exception of meningioma (D32) as an obesity-related cancer.

We defined 13 cancer types as ORCs using the International Agency for Research in Cancer criteria(12).These included: pancreatic (C25), hepatocellular (C22.0), colorectal (C18.0, C18.2-C18.9, C19, and C20), post-menopausal breast (C50), endometrial (C54.1), oesophageal adenocarcinoma ( C15, and ICD-O-3 morphology codes 8140–8141, 8143– 8145, 8190–8231, 8260–8263, 8310, 8401, 8480–8490, 8550–8551, 8570–8574, 8576), kidney (C64), gastric cardia (C16.0), gallbladder (C23), ovarian (C56), thyroid (C73), multiple myeloma (C90.0), and meningioma (D32.0,D32.1,D32.9,C70.0,C70.1,C70.9). We defined post-menopausal breast cancer using a breast cancer diagnosis following the age of self-reported menopause. In the scenario where self-reported menopause age was unavailable, we defined post-menopausal breast cancer using an age cut-off of 55 years. Participants who underwent a bilateral oophorectomy or hysterectomy were excluded from this definition to minimise inclusion of both pre-mature and the medically induced menopause.

We defined NORCs as total cancers minus ORCs. We repeated our analyses excluding prostate cancer due to the well-established inverse association with T2DM(21). Where case number permitted (minimum of 30 cases in both exposed and matched unexposed groups), we further investigated the risk of obesity-related and non-obesity related site-specific cancers.

### Matching

To reduce the confounding effect of obesity, we developed a matched cohort of participants with new-onset T2DM versus participants unexposed at the time of matching (1:3), principally based on matching by BMI. All participants were cancer-free at model enrolment. Participants were matched having a BMI value within 0.1 standard deviations of the exposed participant, within a 5-year age range. Men and women were matched separately. We excluded participants with missing baseline BMI measurements, and those lost to follow-up. To further balance covariate distribution, we used a sequential longitudinal matching algorithm as described by Thomas et al, 2020(16).

We adjusted all cox models for baseline reported values of BMI, Townsend deprivation, summed MET (metabolic equivalent task) minutes per week, smoking status (current, previous, and never), and alcohol status (current, previous, and never).

To address the issue of missingness in our data, we performed multiple imputation by chained equations (MICE) using the mice package in R.(22).Ten imputed datasets were generated using predictive mean matching (PMM) for continuous variables and polytomous logistic regression (polyreg) for categorical variables. The imputation model included all analysis variables (T2DM, BMI, physical activity, smoking habit, alcohol use, Townsend deprivation index, age, and sex), along with the cancer outcome and survival time (23). All missing covariates were imputed for, except BMI due to its central role in the analyses and was therefore restricted to complete cases for this variable. The imputed datasets were analysed and pooled according to Rubin’s rules using the mitools package in R (24).

Sensitivity analyses were performed replacing BMI with waist circumference (WC) and waist-to-hip ratio (WHR).

### Statistical analysis

Time-split hazard ratios (tsHRs) comparing participants with new-onset T2DM with matched unexposed participants were estimated using stratified Cox proportional hazards models, with strata defined by matched group. Follow-up was split at 1 year and an interaction between T2DM and time was included to account for potential detection-time bias and reverse causality. Follow-up began at T2DM diagnosis for exposed participants and the corresponding index date for matched unexposed participants, and ended at first cancer diagnosis, death, or the relevant cancer-registry censoring date, whichever occurred first. Matched unexposed participants who developed T2DM during follow-up were censored at diagnosis and subsequently contributed follow-up as exposed. Models were adjusted for BMI, Townsend deprivation index, physical activity (MET-min/week), smoking status and alcohol consumption, in addition to matching on age, sex and BMI. Proportional hazards assumptions were assessed using Schoenfeld residuals, and Fine–Gray competing-risk models were performed considering death before cancer diagnosis or diagnosis of another primary cancer as competing events.

To examine temporal patterns in cancer incidence, follow-up was additionally divided into predefined intervals and incidence rates calculated separately for exposed and unexposed participants per 100,000 person-years. Incidence rate ratios (IRRs) and 95% confidence intervals were estimated assuming a Poisson distribution. Temporal trends were modelled using Poisson regression with a log person-time offset and restricted cubic splines for time since diagnosis, with separate models for men and women. All tests were two-sided, with P<0.05 considered statistically significant. Analyses were conducted in R version 4.3.1 using the dplyr, magrittr, tidyr, survival, splines, crrSC, mice and mitools packages.

## Results

### New-onset T2DM cohort and matching

The flow diagram from the initial 502,143 participants enrolled in UK Biobank to the matched cohorts is shown in **Figure 1**. After applying our inclusion and exclusion criteria, there were 23,978 participants with new onset T2DM.

**Figure 1.**
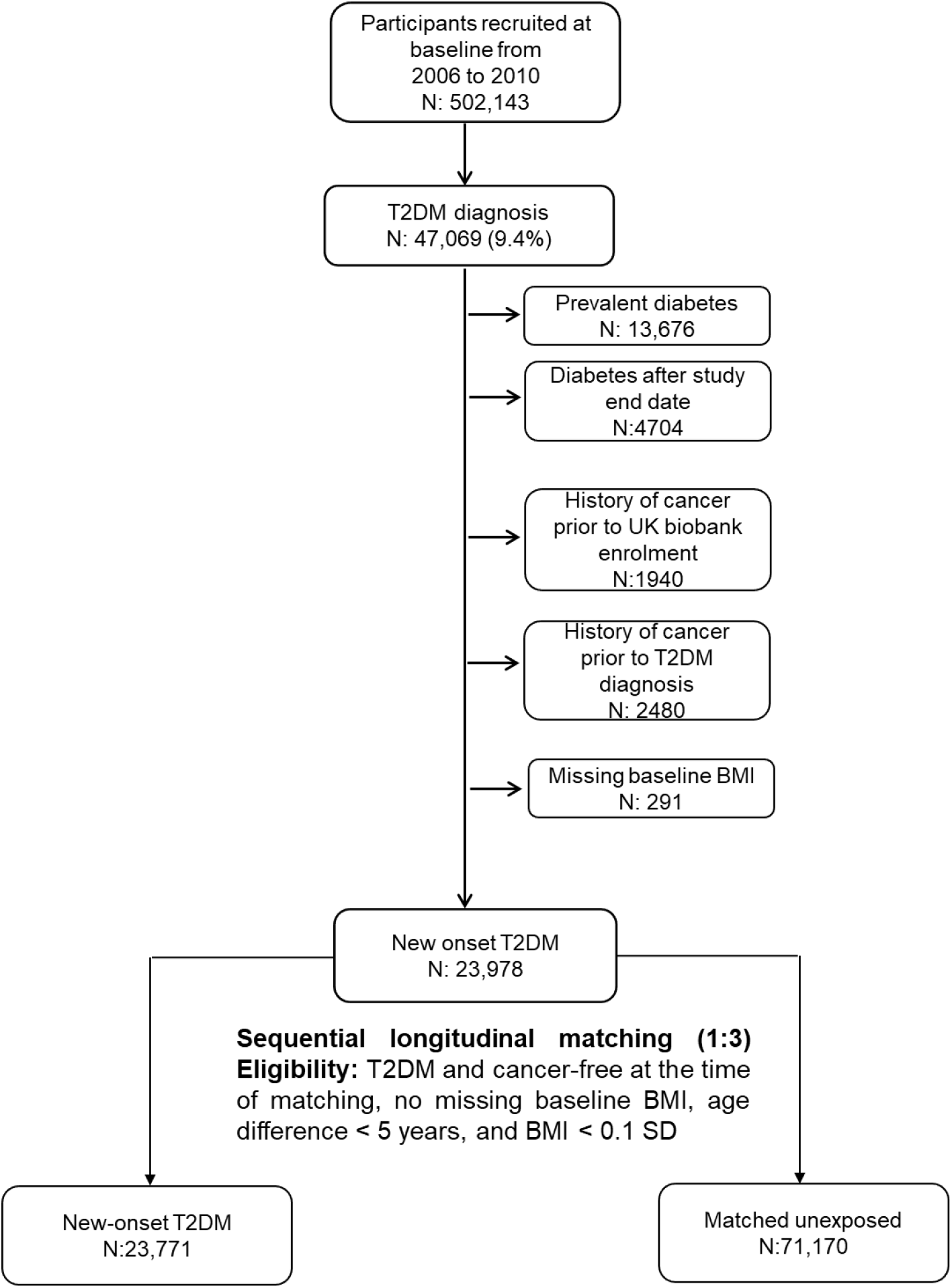
Flow diagram illustrating how new-onset T2DM was defined and the subsequent 1:3 matching. All participants were eligible for matching if they satisfied dynamic matching criteria at the matching time-point of the exposed participant.

Prior to matching, there were significant differences in covariate balance between UK Biobank participants with new-onset T2DM and eligible unexposed participants at baseline (<u>Supplementary Table 1</u>).

The baseline characteristics after matching are shown in **Table 1**. Mean BMI was well matched – for men, the mean BMI values for new-onset T2DM and matched unexposed participants diabetes were 30.90 kg/m^2^ and 30.80 kg/m^2^, respectively. For women, the mean BMI values for new-onset T2DM and never diabetes 32.14 kg/m^2^ and 32.09 kg/m^2^, respectively. Not unexpectedly, participants with new-onset T2DM were more likely to be current smokers and never drinkers, had greater socioeconomic deprivation, and had lower mean physical activity levels in both men and women. During follow-up, approximately 6.92% of unexposed matched men participants developed T2DM, in comparison to 5.98%. Men (n=185) and women (n=22) with new-onset T2DM failed to get a single match likely due to extreme covariate values and were excluded. Unmatched men had a mean BMI of 44.12 kg/m² (SD: 4.71) and a mean age at T2DM diagnosis of 69.42 years (SD:6.37), while unmatched women had a mean BMI of 53.71 kg/m² (SD: 5.74) and a mean age at diagnosis of 64.62 years (SD: 11.36).

**Table 1.** Baseline characteristics of UK Biobank participants post-matching.

|  | Men |  | Women |  |
| --- | --- | --- | --- | --- |
|  | New-onset T2DM | Matched unexposed | New-onset T2DM | Matched unexposed |
| **Number of participants** | 14,009 | 41,911 | 9762 | 29,259 |
| **Mean body mass index (SD) kg/m<sup>2</sup>** | 30.90 (4.88) | 30.80 (4.81) | 32.14 (6.26) | 32.09 (6.21) |
| **Median follow up time in years (IQR)** | 5.03 (2.34-8.00) | 5.10 (2.50-8.03) | 4.88 (2.32-7.71) | 4.99 (2.46-7.8) |
| **Mean follow-up time in years (SD)** | 5.28 (3.43) | 5.37 (3.38) | 5.17 (3.34) | 5.26 (3.34) |
| **Mean Index age (SD)** | 65.2 (7.89) | 64.80 (7.70) | 65.45 (7.99) | 65.09 (7.76) |
| **Smoking (%)** |  |  |  |  |
| Current smoker | 2132 (15.2) | 4409 (10.5) | 1205 (12.3) | 2131 (7.3) |
| Previous smoker | 6438 (46.0) | 18,329 (43.7) | 3073 (31.5) | 9752 (33.3) |
| Never smoker | 5351 (38.2) | 18,988 (45.3) | 5429 (55.6) | 17,247 (58.9) |
| Prefer not to answer | 88 (0.6) | 185 (0.4) | 55 (0.6) | 129 (0.4) |
| **Alcohol consumption (%)** |  |  |  |  |
| Current alcohol drinker | 12,474 (89.0) | 39,379 (94.0) | 7819 (80.1) | 25,984 (88.8) |
| Previous alcohol drinker | 774 (5.5) | 1435 (3.4) | 677 (6.9) | 1231 (4.2) |
| Never alcohol drinker | 724 (5.2) | 1047 (2.5) | 1249 (12.8) | 1993 (6.8) |
| Prefer not to answer | 37 (0.3) | 50 (0.1) | 17 (0.2) | 51 (0.2) |
| **Mean Townsend deprivation index (SD)** | -0.462 (3.48) | -1.30 (3.12) | -0.304 (3.39) | -1.10 (3.14) |
| **Mean summed MET minutes (SD)** | 2482 (2795) | 2629 (2750) | 2315 (2516) | 2357 (2443) |

SD: standard deviation. IQR: inter-quartile range. MET: metabolic equivalent task.

### Cancer incidence in new-onset type 2 diabetes mellitus

With a median follow-up of 5.0 years, there were 2432 (men: 1590; women: 842) new cancers among participants with new-onset T2DM and 5262 (men: 3553; women: 1709) new cancers among matched unexposed participants. In men, the unadjusted incidence per 100,000 person years for total cancer was 2150 (95% CI: 2046-2258) in participants with new-onset diabetes compared with 1579 (95% CI: 1528-1632) for matched unexposed participants. Once prostate cancer was excluded from the outcome set, men with new-onset T2DM had a total cancer incidence of 1605 (95% CI:1515-1699) per 100,000 person years compared with 952 (95% CI: 912-993) for matched unexposed participants. For women, the unadjusted incidence per 100,000 person years for total cancer was 1668 (95% CI: 1557-1784) in participants with new-onset diabetes compared with 1110 (95% CI: 1058-1164) for matched unexposed participants.

### Evidence of detection time bias

In men, cancer incidence rates were greater with new-onset T2DM during the first year of follow-up after diagnosis of new-onset T2DM for all cancer, ORC, and NORC, compared to the follow-up period beyond 1 year **(Figure 2)**. In women, we observed an increased occurrence of cancer within the first year of diagnosis, but to a lesser extent than in men. These observations justified modelling and reporting time-split hazard ratios (tsHRs).

**Figure 2.**
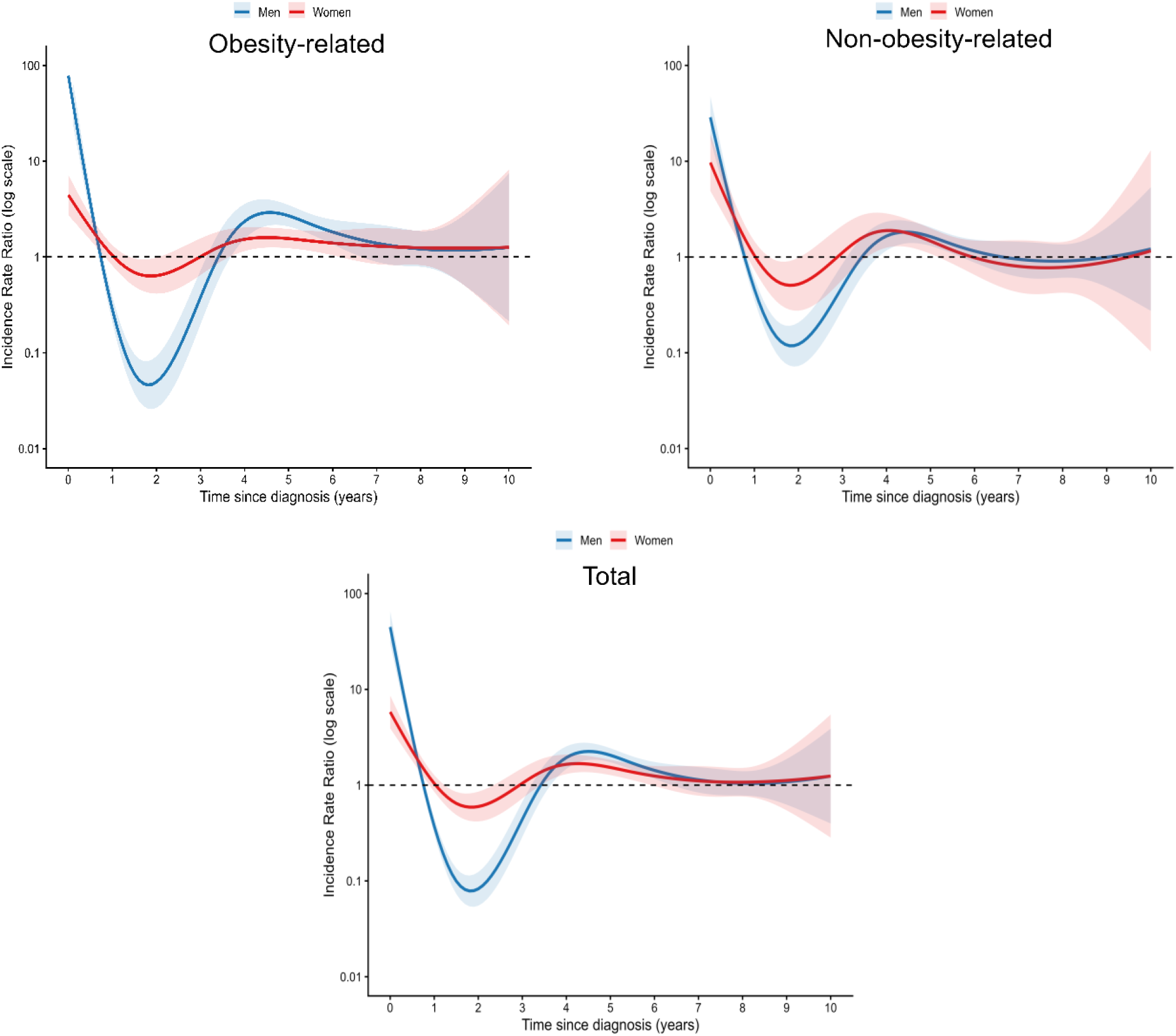
Incidence rate ratios (IRRs) for cancer over time since diagnosis estimated using a Poisson regression model with restricted cubic splines. Separate models were fitted for men and women, with a log person-time offset to account for follow-up time. IRRs are shown on a logarithmic scale with 95% CIs derived from the model. The dashed line indicates an IRR of 1, and shaded areas represent 95% confidence intervals. This plot was restricted to a maximum of 10 years due to low cancer events and event-free progression past this time-point.

### Associations with risk of total cancer, ORC and NORC

There was evidence for an effect of new-onset T2DM on total cancer in women (tsHR 1.32, 95% CI 1.19-1.46), but limited evidence for an effect in men (tsHR 1.06, 95% CI 0.98-1.14) as shown in **Table 2**. However, after excluding prostate cancer, there was evidence for an effect on total cancer in men (tsHR 1.21, 95% CI 1.10-1.32). There was evidence for an effect of new-onset T2DM on ORCs in men (tsHR 1.39, 95% CI 1.21-1.59), and for an effect in women (tsHR 1.33, 95% CI 1.16-1.52). There was no effect of new-onset T2DM on NORCs in men and women. When prostate cancers were excluded from the outcome set in men, evidence for no effect on NORCs remained.

**Table 2.** Times-split hazard ratios (tsHRs) and 95% confidence intervals (CIs) for cancer outcomes in men and women with new-onset type 2 diabetes mellitus (T2DM)

|  | Men |  | Women |  |
| --- | --- | --- | --- | --- |
|  | New-onset T2DM | Matched unexposed | New-onset T2DM | Matched unexposed |
| Participants | 14,009 | 41,911 | 9762 | 29,259 |
| Person years | 73,951 | 224,970 | 50,484 | 153,943 |
| **TOTAL CANCER** |  |  |  |  |
| No. of cancers | 1187 | 2141 | 842 | 1709 |
| tsHR (95%CIs) | 1.21 (1.10-1.32) | 1 (Reference) | 1.32 (1.19-1.46) | 1 (Reference) |
| **OBESITY-RELATED CANCERS** |  |  |  |  |
| No. of cancers | 561 | 889 | 487 | 1003 |
| tsHR (95%CIs) | 1.39 (1.21-1.59) | 1 (Reference) | 1.33 (1.16-1.52) | 1 (Reference) |
| **Liver** |  |  |  |  |
| No. of cancers | 71 | 38 | 13 | 9 |
| tsHR (95%CIs) | 3.97 (2.38-6.65) | 1 (Reference) | Not estimated | 1 (Reference) |
| **Pancreas** |  |  |  |  |
| No. of cancers | 100 | 84 | 64 | 60 |
| tsHR (95%CIs) | 1.77 (1.15-2.72) | 1 (Reference) | 1.63 (0.99-2.70) | 1 (Reference) |
| **Colorectal** |  |  |  |  |
| No. of cancers | 209 | 391 | 83 | 183 |
| tsHR (95%CIs) | 1.22 (0.99-1.51) | 1 (Reference) | 1.18 (0.85-1.62) | 1 (Reference) |
| **Post-menopausal breast cancer** |  |  |  |  |
| No. of cancers |  |  | 127 | 381 |
| tsHR (95%CIs) |  |  | 1.18 (0.93-1.49) | 1 (Reference) |
| **Endometrial** |  |  |  |  |
| No. of cancers |  |  | 78 | 176 |
| tsHR (95%CIs) |  |  | 1.17 (0.83-1.65) | 1 (Reference) |
| **Ovarian** |  |  |  |  |
| No. of cancers |  |  | 26 | 62 |
| tsHR (95%CIs) |  |  | Not estimated | 1 (Reference) |
| **Kidney** |  |  |  |  |
| No. of cancers | 82 | 127 | 24 | 39 |
| tsHR (95%CIs) | 1.62 (1.13-2.32) | 1 (Reference) | Not estimated | 1 (Reference) |
| **Oesophageal** |  |  |  |  |
| No. of cancers | 33 | 83 | 9 | 7 |
| tsHR (95%CIs) | 0.99 (0.59-1.68) | 1 (Reference) | Not estimated | 1 (Reference) |
| **Gastric cardia** |  |  |  |  |
| No. of cancers | 14 | 37 | 5 | 4 |
| tsHR (95%CIs) | Not estimated | 1 (Reference) | Not estimated | 1 (Reference) |
| **Multiple myeloma** |  |  |  |  |
| No. of cancers | 32 | 88 | 17 | 24 |
| tsHR (95%CI) | 0.78 (0.46-1.32) | 1 (Reference) | Not estimated | 1 (Reference) |
| **Gallbladder** |  |  |  |  |
| No. of cancers | 3 | 7 | 6 | 8 |
| tsHR (95%CI) | Not estimated | 1 (Reference) | Not estimated | 1 (Reference) |
| **Thyroid** |  |  |  |  |
| No. of cancers | 6 | 16 | 19 | 23 |
| tsHR (95%CI) | Not estimated | 1 (Reference) | Not estimated | 1 (Reference) |
| **Meningioma** |  |  |  |  |
| No. of cancers | 5 | 13 | 16 | 27 |
| tsHR (95%CI) | Not estimated | 1 (Reference) | Not estimated | 1 (Reference) |
| **NON-OBESITY-RELATED CANCERS** |  |  |  |  |
| No. of cancers | 626 | 1252 | 355 | 706 |
| tsHR (95%CI) | 1.07 (0.95-1.22) | 1 (Reference) | 1.03 (0.88-1.21) | 1 (Reference) |
| **Prostate** |  |  |  |  |
| No. of cancers | 403 | 1412 |  |  |
| tsHR (95%CI) | 0.81 (0.71-0.93) | 1 (Reference) |  |  |
| **Lung** |  |  |  |  |
| No. of cancers | 157 | 257 | 97 | 134 |
| tsHR (95%CI) | 1.11 (0.83-1.48) | 1 (Reference) | 1.63 (1.10-2.43) | 1 (Reference) |
| **Bladder** |  |  |  |  |
| No. of cancers | 66 | 100 | 13 | 19 |
| tsHR (95%CI) | 1.36 (0.91-2.04) | 1 (Reference) | Not estimated | 1 (Reference) |
| **Melanoma** |  |  |  |  |
| No. of cancers | 50 | 181 | 23 | 85 |
| tsHR (95%CI) | 0.98 (0.68-1.41) | 1 (Reference) | Not estimated | 1 (Reference) |
| **Non-follicular lymphoma** |  |  |  |  |
| No. of cancers | 40 | 85 | 12 | 38 |
| tsHR (95%CI) | 1.50 (0.91-2.46) | 1 (Reference) | Not estimated | 1 (Reference) |

Missing data were handled using multiple imputation (10 datasets). Matching was performed on BMI (0.1 SD calliper) and age (<5-year interval), with stratification by matched set. Cox models were additionally adjusted for BMI, smoking, alcohol, Townsend deprivation index, and total physical activity (MET minutes/week), and included a T2DM × time interaction. Estimates were pooled using Rubin's rules. In men, total and non-obesity-related cancer estimates excluded prostate cancer

### Associations with risk for site-specific cancer types

In men, there was evidence for site-specific effects on hepatocellular carcinoma (tsHR 3.97, 95% CI 2.38-6.65), pancreatic cancer (tsHR 1.77, 95% CI 1.15-2.72), kidney cancer (tsHR 1.53, 95% CI 1.08-2.17), and weak evidence for colorectal cancer (tsHR 1.22, 95% CI 0.99-1.51). There was evidence of no site-specific associations in men with oesophageal adenocarcinoma and multiple myeloma. The remaining ORCs did not have sufficient cases for site-specific analysis.

In women, there was weak evidence for a site-specific effect on pancreatic cancer (tsHR 1.63, 95% CI 0.99-2.70). There was evidence for no effect on endometrial, post-menopausal breast, and colorectal cancers. The remaining ORCs did not have sufficient cases for site-specific analysis.

When investigating site-specific NORCs, there was evidence of an association with lung cancer in women (tsHR 1.63, 95% CI 1.10–2.43), but not in men. In men, there was weak evidence of an association with bladder cancer (tsHR 1.36, 95% CI 0.91–2.04) and non-follicular lymphoma (tsHR 1.50, 95% CI 0.91–2.46), whereas prostate cancer risk was reduced (tsHR 0.81, 95% CI 0.71–0.93). In women, case numbers were insufficient to reliably assess associations with bladder cancer and non-follicular lymphoma.

### Sensitivity analyses

To assess the impact of detection bias/reverse causality, estimates obtained before and after applying the 1-year time split were compared to observe whether effect estimates were inflated during the first year of follow up. In men, estimates were significantly inflated within the first year for total cancer, ORC, and NORC, whilst in women, estimates did not show significant inflation as shown in **Figure 3**.

**Figure 3.**
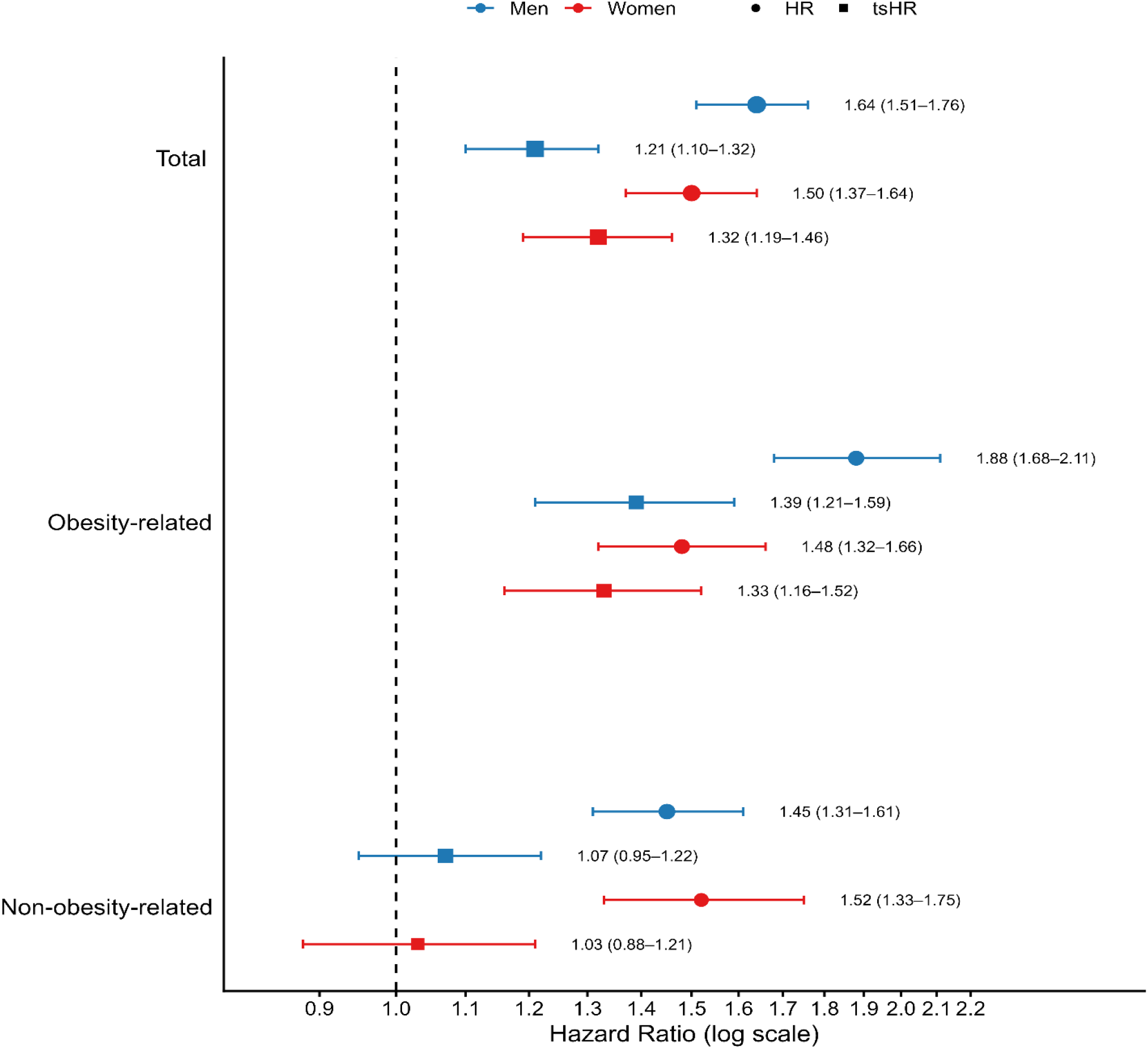
Coefficient plot illustrating HRs and tsHRs with 95% CIs for the association between new-onset T2DM and cancer outcomes in men and women, before and after applying a 1-year time split. Estimates for total cancers and NORCs in men exclude prostate cancer.

When the proportional hazards were assessed using Schoenfeld residuals without time-splitting, evidence of violation was observed (<u>Supplementary Table 19</u>). After applying the time-split, the proportional hazards assumption was no longer violated (<u>Supplementary Table 20</u>).

Results were also robust to alternative adiposity measures, with analyses using WC and WHR yielding estimates that were consistent in direction and magnitude (<u>Supplementary Tables 3– 11</u>). Estimates from Fine–Gray competing risks models were also comparable to those from the main Cox models (<u>Supplementary Tables 12-14</u>).

## Discussion

### Main findings

After controlling for the key confounding of obesity, first, we found associations between new-onset T2DM and ORCs limited to a small number of cancer types, namely with pancreatic, liver, and kidney cancer in men and weak evidence of association with pancreatic cancer in women. Second, there was evidence for no associations with endometrial and post-menopausal breast cancers, both of which have been previously linked to T2DM (25–27). Third, there was evidence for no association between T2DM and NORCs, with two exceptions: the well-known inverse effect with prostate cancer in men, and an unexpected significant effect on lung cancer in women, which might be attributed to residual confounding. Fourth, we demonstrated that these effects were consistent when approximates of obesity other than BMI, namely WC and WHR, were evaluated. Finally, we found clear evidence for detection time bias and if this is not accounted for, risk estimates of associations between T2DM and cancer are overinflated. Taken together, these observations suggest that T2DM may be causally associated with a small number of ORCs, namely pancreatic and probably liver, independent of obesity pathways.

### Literature context

The most notable finding in this study was the association with pancreatic cancer, present in men and women, and aligned with findings in previous observational and Mendelian Randomisation studies (<u>Supplementary Table 24</u>) (3,4). There was also strong evidence for an effect on hepatocellular carcinoma in men, but this finding was not replicated in women, due to limited case numbers. The lack of MR studies investigating T2DM and hepatocellular carcinoma, due to its low incidence in Western populations, limits the ability to triangulate evidence and leads to some uncertainty regarding the causal nature of this association(28). Previously, associations with pancreatic and hepatocellular carcinoma have been thought to reflect reverse causality, rather than a true causal link (29). Despite this, after adjustment for these biases, the effect estimate remained significant. There was also evidence for associations with kidney cancer in men, and a possible effect on colorectal cancer in men, which remains consistent with observational research (30, 31). Moreover, there was also evidence for an inverse effect on prostate cancer in men, and a positive effect on lung cancer in women. Upon triangulation with MR studies however, these findings remain limited in supporting a causal effect for colorectal, prostate, and lung cancer (3,4).

This study did not support an association with endometrial, or post-menopausal breast cancers, which contradicts previous observational research. Overall, our results upon triangulation with MR studies suggest that T2DM may be causally associated with some but not all ORCs, primarily pancreatic cancer, and probably hepatocellular carcinoma. Moreover, the lack of evidence for an effect on endometrial and post-menopausal breast cancer in women could suggest that previously established T2DM-related cancers in observational research may be driven through confounding obesity and detection-time bias. Furthermore, these results have been closely replicated using identical methodology in the China Kadoorie Biobank (CKB), which demonstrated significant associations with digestive accessory organ cancers (mainly, liver and pancreas), but an absence of any effect on female reproductive organ cancers (endometrial, ovarian, and post-menopausal breast) (32).

### Strengths and limitations

We adjusted for detection time bias, reverse causality, and attempted to eliminate the effects of potential confounding due to BMI through matching and adjustment in our Cox models. Moreover, our application of sequential longitudinal matching enabled us to match on BMI, age, and sex using a strict calliper, and handle unexposed participants who subsequently developed T2DM during follow up, which was likely due to matching on BMI, a risk factor for T2DM. Similarly, using new-onset T2DM as the exposure and excluding prevalent T2DM helped to reduce the effect of immortal-time bias.

There are also limitations. Mainly, from reduced sample numbers after matching, we suffered from limited case numbers and weak statistical power when investigating several site-specific cancers. With the innate design of UK Biobank, we acknowledge that there will be some element of selection bias within participant recruitment, which may affect the generalizability of our estimates to the wider UK population(33). Similarly, we acknowledge the heterogeneous nature of obesity, and limitation of using baseline BMI as an imperfect surrogate marker, which may not capture the life-course effects of excess adiposity.

### Unanswered questions and future research

For several cancer types in our study, it remains unclear whether there is an association between T2DM and site-specific cancers due to low cancer numbers. Furthermore, whilst we identified several associations between new-onset T2DM and cancer, replication of this research in alternative European cohorts is required to verify the robustness of these findings. Our findings show strong evidence for an association between new-onset T2DM and ORCs, primarily driven through pancreatic cancer in men and women, and liver and kidney cancer in men. Triangulation from MR studies provides evidence to suggest a causal relationship between T2DM and pancreatic cancer and possibly kidney cancer(1, 34). However, for hepatocellular carcinoma, the causal nature of this association remains unknown due to the lack of genome-wide association studies (GWAS) for hepatocellular carcinoma in European ancestries.

### Conclusion

We found evidence that T2DM may be causally associated with a small number of ORCs, namely pancreatic and probably liver, independent of obesity pathways. We also found clear evidence for detection time bias and if this is not accounted for, risk estimates of associations between T2DM and cancer are overinflated. We urge researchers to adequately adjust for obesity parameters, and detection-time bias when investigating T2DM-related cancer risk in an observational setting to provide more robust estimates, and champion the idea of triangulation to help tackle the issue of bias within observational research.

## Data and resource availability

Researchers can apply to use the UK Biobank resource, and access all data used within this study. This research has been conducted using the UK Biobank Resource under Application Number 196956.

## Contributors

OT: Conceptualization, methodology development, data analysis, writing-original manuscript. MW, RM,MS: Conceptualization, methodology development, writing-review/editing. AGR: Conceptualisation, project administration, funding acquisition, writing-review /editing.

## Acknowledgements

AGR receives funding through the NIHR Manchester Biomedical Research Centre (BRC) (NIHR203308). Cancer Research UK. R.M.M. is a National Institute for Health Research Senior Investigator (NIHR202411). R.M.M. is supported by a Cancer Research UK 25 (C18281/A29019) programme grant (the Integrative Cancer Epidemiology Programme). R.M.M. is also supported by the NIHR Bristol Biomedical Research Centre which is funded by the NIHR (BRC-1215-20011) and is a partnership between University Hospitals Bristol and Weston NHS Foundation Trust and the University of Bristol. MS acknowledges support of the UKRI AI programme, and the Engineering and Physical Sciences Research council, for CHAI – Causality in Healthcare AI Hub [grant number EP/Y028856/1]

## References

1. Pearson-Stuttard J, Papadimitriou N, Markozannes G, Cividini S, Kakourou A, Gill D, et al. Type 2 Diabetes and Cancer: An Umbrella Review of Observational and Mendelian Randomization Studies. Cancer Epidemiol Biomarkers Prev. 2021;30(6):1218–28.

2. Tsilidis KK, Kasimis JC, Lopez DS, Ntzani EE, Ioannidis JP. Type 2 diabetes and cancer: umbrella review of meta-analyses of observational studies. BMJ. 2015;350:g7607.

3. Ling S, Brown K, Miksza JK, Howells L, Morrison A, Issa E, et al. Association of Type 2 Diabetes With Cancer: A Meta-analysis With Bias Analysis for Unmeasured Confounding in 151 Cohorts Comprising 32 Million People. Diabetes Care. 2020;43(9):2313–22.

4. Starup-Linde J, Karlstad O, Eriksen SA, Vestergaard P, Bronsveld HK, de Vries F, et al. CARING (CAncer Risk and INsulin analoGues): the association of diabetes mellitus and cancer risk with focus on possible determinants - a systematic review and a meta-analysis. Curr Drug Saf. 2013;8(5):296–332.

5. Noto H, Osame K, Sasazuki T, Noda M. Substantially increased risk of cancer in patients with diabetes mellitus: a systematic review and meta-analysis of epidemiologic evidence in Japan. J Diabetes Complications. 2010;24(5):345–53.

6. Ohkuma T, Peters SAE, Woodward M. Sex differences in the association between diabetes and cancer: a systematic review and meta-analysis of 121 cohorts including 20 million individuals and one million events. Diabetologia. 2018;61(10):2140–54.

7. De Bruijn KM, Arends LR, Hansen BE, Leeflang S, Ruiter R, van Eijck CH. Systematic review and meta-analysis of the association between diabetes mellitus and incidence and mortality in breast and colorectal cancer. Br J Surg. 2013;100(11):1421–9.

8. Wu W, Huang G-l, Cui J. Type 2 diabetes mellitus and cancer: A systematic review and meta-analysis of Mendelian randomization studies. Frontiers in Endocrinology. 2026;Volume 17 – 2026.

9. Pearson-Stuttard J, Zhou B, Kontis V, Bentham J, Gunter MJ, Ezzati M. Worldwide burden of cancer attributable to diabetes and high body-mass index: a comparative risk assessment. Lancet Diabetes Endocrinol. 2018;6(6):e6–e15.

10. Pearson-Stuttard J, Buckley J, Cicek M, Gregg EW. The Changing Nature of Mortality and Morbidity in Patients with Diabetes. Endocrinol Metab Clin North Am. 2021;50(3):357–68.

11. Renehan AG, Tipping O, Wang M. Diabetes and cancer: Doubts of a causal link. Int J Cancer. 2024;154(11):1875–6.

12. Lauby-Secretan B, Scoccianti C, Loomis D, Grosse Y, Bianchini F, Straif K, et al. Body Fatness and Cancer–Viewpoint of the IARC Working Group. N Engl J Med. 2016;375(8):794–8.

13. Cohen DH, LeRoith D. Obesity, type 2 diabetes, and cancer: the insulin and IGF connection. Endocrine-related cancer. 2012;19(5):F27–F45.

14. Joung KH, Jeong JW, Ku BJ. The association between type 2 diabetes mellitus and women cancer: the epidemiological evidences and putative mechanisms. Biomed Res Int. 2015;2015:920618.

15. Zhang Z, Reinikainen J, Adeleke KA, Pieterse ME, Groothuis-Oudshoorn CGM. Time-varying covariates and coefficients in Cox regression models. Ann Transl Med. 2018;6(7):121.

16. Thomas LE, Yang S, Wojdyla D, Schaubel DE. Matching with time-dependent treatments: A review and look forward. Stat Med. 2020;39(17):2350–70.

17. Ballotari P, Vicentini M, Manicardi V, Gallo M, Chiatamone Ranieri S, Greci M, et al. Diabetes and risk of cancer incidence: results from a population-based cohort study in northern Italy. BMC Cancer. 2017;17(1):703.

18. Hernan MA, Sauer BC, Hernandez-Diaz S, Platt R, Shrier I. Specifying a target trial prevents immortal time bias and other self-inflicted injuries in observational analyses. J Clin Epidemiol. 2016;79:70–5.

19. Harding JL, Shaw JE, Peeters A, Cartensen B, Magliano DJ. Cancer risk among people with type 1 and type 2 diabetes: disentangling true associations, detection bias, and reverse causation. Diabetes Care. 2015;38(2):264–70.

20. Sudlow C, Gallacher J, Allen N, Beral V, Burton P, Danesh J, et al. UK biobank: an open access resource for identifying the causes of a wide range of complex diseases of middle and old age. PloS Med. 2015;12(3):e1001779.

21. Bansal D, Bhansali A, Kapil G, Undela K, Tiwari P. Type 2 diabetes and risk of prostate cancer: a meta-analysis of observational studies. Prostate cancer and prostatic diseases. 2013;16(2):151–8.

22. van Buuren S, Groothuis-Oudshoorn K. mice: Multivariate Imputation by Chained Equations in R. Journal of Statistical Software. 2011;45(3):1 –67.

23. Austin PC, White IR, Lee DS, van Buuren S. Missing Data in Clinical Research: A Tutorial on Multiple Imputation. Can J Cardiol. 2021;37(9):1322–31.

24. Lumley T. mitools: Tools for multiple imputation of missing data. URL http://CRAN R- project org. 2006.

25. Tsilidis KK, Kasimis JC, Lopez DS, Ntzani EE, Ioannidis JPA. Type 2 diabetes and cancer: umbrella review of meta-analyses of observational studies. BMJ. 2015;350(jan02 1):g7607-g.

26. Friberg E, Orsini N, Mantzoros CS, Wolk A. Diabetes mellitus and risk of endometrial cancer: a meta-analysis. Diabetologia. 2007;50(7):1365–74.

27. Larsson SC, Mantzoros CS, Wolk A. Diabetes mellitus and risk of breast cancer: a meta-analysis. Int J Cancer. 2007;121(4):856–62.

28. McGlynn KA, London WT. The global epidemiology of hepatocellular carcinoma: present and future. Clin Liver Dis. 2011;15(2):223–43, vii-x.

29. Johnson JA, Carstensen B, Witte D, Bowker SL, Lipscombe L, Renehan AG. Diabetes and cancer (1): evaluating the temporal relationship between type 2 diabetes and cancer incidence. Diabetologia. 2012;55(6):1607–18.

30. Larsson SC, Wolk A. Diabetes mellitus and incidence of kidney cancer: a meta-analysis of cohort studies. Diabetologia. 2011;54(5):1013–8.

31. Larsson SC, Orsini N, Wolk A. Diabetes mellitus and risk of colorectal cancer: a meta-analysis. J Natl Cancer Inst. 2005;97(22):1679–87.

32. Wang M, Tipping O, Liu B, Bennett DA, Shen B, Zou S, et al. New-Onset Type 2 Diabetes Mellitus and Cancer Risk: A Matched Cohort Study in China Kadoorie Biobank. Int J Cancer. 2026.

33. Fry A, Littlejohns TJ, Sudlow C, Doherty N, Adamska L, Sprosen T, et al. Comparison of Sociodemographic and Health-Related Characteristics of UK Biobank Participants With Those of the General Population. Am J Epidemiol. 2017;186(9):1026–34.

34. Wu W, Huang G-l, Cui J. Type 2 diabetes mellitus and cancer: A systematic review and meta-analysis of Mendelian randomization studies. Frontiers in Endocrinology. 2026;17:1713815.

