## Supplementary materials for "New-onset type 2 diabetes mellitus and obesity-related cancer risk: a matched cohort study (UK Biobank)"

**Supplementary Table 1 Characteristics of participants with new-onset T2DM and never diabetes participants prior to matching.**

| | Men | | Women | |
|---|---|---|---|---|
| | New-onset T2DM | Never diabetes | New-onset T2DM | Never diabetes |
| **Number of participants** | 14,194 | 191,050 | 9784 | 232,922 |
| **Mean body mass index (SD) kg/m$^2$** | 31.05 (5.10) | 27.41 (3.92) | 32.91 (6.34) | 26.71 (4.89) |
| **Mean age at recruitment** | 59.39 (7.44) | 56.65 (8.25) | 59.27 (7.45) | 56.43 (8.05) |
| **Smoking (%)** | | | | |
| Current smoker | 2146 (15.12) | 23,552 (12.33) | 1202 (12.29) | 20,009 (8.59) |
| Previous smoker | 6527 (45.98) | 70,231 (36.76) | 3075 (31.43) | 71,611 (30.74) |
| Never smoker | 5404 (38.07) | 96,369 (50.44) | 5432 (55.52) | 140,252 (60.21) |
| Prefer not to answer | 89 (0.63) | 716 (0.37) | 55 (0.56) | 824 (0.35) |
| Missingness | 28 (0.20) | 182 (0.09) | 20 (0.20) | 226 (0.10) |
| **Alcohol consumption (%)** | | | | |
| Current alcohol drinker | 12,621 (88.92) | 180,022 (94.23) | 7820 (79.93) | 212,391 (91.19) |
| Previous alcohol drinker | 781 (5.50) | 5931 (3.10) | 677 (6.92) | 7667 (3.29) |
| Never alcohol drinker | 726 (5.11) | 4645 (2.43) | 1250 (12.77) | 12,351 (5.30) |
| Prefer not to answer | 38 (0.27) | 269 (0.14) | 17 (0.17) | 285 (0.12) |
| Missingness | 28 (0.20) | 183 (0.09) | 20 (0.20) | 228 (0.10) |
| **Mean Townsend deprivation index (SD)** | -0.452 (3.47) | -1.36 (3.10) | -0.305 (3.39) | -1.41 (3.00) |
| Missingness | 21 (0.15) | 251 (0.13) | 18 (0.18) | 262 (0.11) |
| **Mean summed MET minutes (SD)** | 2466 (2850) | 2825 (2865) | 2263 (2462) | 2555 (2450) |
| Missingness | 3637 (25.62) | 35,290 (18.47) | 3475 (33.52) | 59,915 (25.72) |

**Supplementary Table 2 International Classification of Diseases, Tenth Revision (ICD-10) coding for all 13 obesity-related cancers (ORCs).**

| Cancer | ICD-10 codes | ORCs | |
|---|---|---|---|
| | | Men | Women |
| All malignant neoplasms | C00–C97 | | |
| Oesophageal adenocarcinoma | C15+ ICD-O-3 morphology codes: 8140–8141, 8143–8145, 8190–8231, 8260–8263, 8310, 8401, 8480–8490, 8550–8551, 8570–8574, 8576 | ● | ● |
| Gastric cardia | C16.0 | ● | ● |
| Colorectal | C18-C20 | ● | ● |
| Hepatocellular carcinoma | C22.0 | ● | ● |
| Gallbladder | C23 | ● | ● |
| Pancreatic | C25 | ● | ● |
| Breast | C50 | | ● |
| Endometrial | C54.1 | | ● |
| Ovarian | C56 | | ● |
| Kidney | C64 | ● | ● |
| Thyroid | C73 | ● | ● |
| Multiple myeloma | C90.1 | ● | ● |
| Meningioma | D32.0, D32.1, D32.9, C70.0, C70.1, C70.9 | ● | ● |

**Supplementary Table** Error! No text of specified style in document. **Pooled hazard ratios (HRs) and 95% confidence intervals (CIs) for obesity-related cancer (ORC) outcomes in men and women with new-onset type 2 diabetes mellitus (T2DM) after performing multiple imputation for missing data and generating 10 imputed datasets, matching on BMI with a 0.1 standard deviation calliper, on age with an interval <5 years, and stratifying on matched set.** Cox model 1a is adjusted for BMI, smoking, alcohol, Townsend deprivation, and Summed Metabolic Equivalent Task (MET) minutes per week for all activity by including them as covariates. Cox model 1b adjusted similarly, but also time-split at 1 year with an interaction term added between T2DM and time, to account for detection-time bias and reverse causality.

| | | **Men** | | **Women** | |
|---|---|---|---|---|---|
| | | **New-onset T2DM** | **Matched controls** | **New-onset T2DM** | **Matched controls** |
| Participants | | 14,009 | 41,911 | 9762 | 29,259 |
| Person-years | | 73,951 | 224,970 | 50,484 | 153,943 |
| ***ORC*** | | | | | |
| Cases | | 561 | 889 | 487 | 1003 |
| IR | | 759 (697-824) | 395 (370-422) | 965 (881-1054) | 652 (612-693) |
| HR (95% CI) | Model 1a | 1.88 (1.68-2.11) | 1 (Reference) | 1.48 (1.32-1.66) | 1 (Reference) |
| | Model 1b | 1.39 (1.21-1.59) | 1 (Reference) | 1.33 (1.16-1.52) | 1 (Reference) |
| ***Liver*** | | | | | |
| Cases | | 71 | 38 | 13 | 9 |
| IR | | 96 (75-121) | 17 (12-23) | 26 (14-44) | 6 (3-11) |
| HR (95% CI) | Model 1a | 4.90 (3.08-7.81) | 1 (Reference) | | 1 (Reference) |
| | Model 1b | 3.97 (2.38-6.65) | 1 (Reference) | | 1 (Reference) |
| ***Pancreas*** | | | | | |
| Cases | | 100 | 84 | 64 | 60 |
| IR | | 135 (110-164) | 37 (30-46) | 48 (30-71) | 39 (30-50) |
| HR (95% CI) | Model 1a | 3.75 (2.73-5.14) | 1 (Reference) | 3.14 (2.12-4.64) | 1 (Reference) |
| | Model 1b | 1.77 (1.15-2.72) | 1 (Reference) | 1.63 (0.99-2.70) | 1 (Reference) |
| ***Colorectum*** | | | | | |
| Cases | | 209 | 391 | 83 | 183 |
| IR | | 283 (246-324) | 174 (157-192) | 164 (131-204) | 119 (102-137) |
| HR (95% CI) | Model 1a | 1.60 (1.33-1.91) | 1 (Reference) | 1.43 (1.08-1.89) | 1 (Reference) |
| | Model 1b | 1.22 (0.99-1.51) | 1 (Reference) | 1.18 (0.85-1.62) | 1 (Reference) |
| ***Postmenopausal breast*** | | | | | |
| Cases | | | | 127 | 381 |
| IR | | | | 252 (210-299) | 247 (223-274) |
| HR (95% CI) | Model 1a | | | 1.01 (0.82-1.25) | 1 (Reference) |
| | Model 1b | | | 1.18 (0.93-1.49) | 1 (Reference) |
| ***Endometrium*** | | | | | |
| Cases | | | | 78 | 176 |
| IR | | | | 155 (122-193) | 114 (98-133) |
| HR (95% CI) | Model 1a | | | 1.40 (1.05-1.86) | 1 (Reference) |
| | Model 1b | | | 1.17 (0.83-1.65) | 1 (Reference) |
| ***Kidney*** | | | | | |
| Cases | | 82 | 127 | 24 | 39 |

| | | | | | |
|---|---|---|---|---|---|
| IR | | 111 (88-138) | 56 (47-67) | 48 (30-71) | 25 (18-35) |
| HR (95% CI) | Model 1a | 2.04 (1.50-2.78) | 1 (Reference) | | 1 (Reference) |
| | Model 1b | 1.62 (1.13-2.32) | 1 (Reference) | | 1 (Reference) |
| ***Oesophagus*** | | | | | |
| Cases | | 33 | 83 | 9 | 7 |
| IR | | 45 (31-63) | 37 (29-46) | 18 (8-34) | 5 (2-9) |
| HR (95% CI) | Model 1a | 1.27 (0.81-1.99) | 1 (Reference) | | 1 (Reference) |
| | Model 1b | 0.99 (0.59-1.68) | 1 (Reference) | | 1 (Reference) |
| ***Gastric cardia*** | | | | | |
| Cases | | 14 | 37 | 5 | 4 |
| IR | | 19 (10-32) | 16 (12-23) | 10 (3-23) | 3 (1-7) |
| HR (95% CI) | Model 1a | | 1 (Reference) | | 1 (Reference) |
| | Model 1b | | 1 (Reference) | | 1 (Reference) |
| ***Multiple myeloma*** | | | | | |
| Cases | | 32 | 88 | 17 | 24 |
| IR | | 43 (30-61) | 39 (31-48) | 34 (20-54) | 16 (10-23) |
| HR (95% CI) | Model 1a | 1.03 (0.66-1.61) | 1 (Reference) | | 1 (Reference) |
| | Model 1b | 0.78 (0.46-1.32) | 1 (Reference) | | 1 (Reference) |
| ***Gallbladder*** | | | | | |
| Cases | | 3 | 7 | 6 | 8 |
| IR | | 4 (1-12) | 3 (1-6) | 12 (4-26) | 5 (2-9) |
| HR (95% CI) | Model 1a | | 1 (Reference) | | 1 (Reference) |
| | Model 1b | | 1 (Reference) | | 1 (Reference) |
| ***Thyroid*** | | | | | |
| Cases | | 6 | 16 | 19 | 23 |
| IR | | 8 (3-18) | 7 (4-12) | 38 (23-59) | 15 (9-21) |
| HR (95% CI) | Model 1a | | 1 (Reference) | | 1 (Reference) |
| | Model 1b | | 1 (Reference) | | 1 (Reference) |
| ***Meningioma*** | | | | | |
| Cases | | 5 | 13 | 16 | 27 |
| IR | | 7(2-16) | 6 (3-10) | 32 (18-51) | 18 (11-24) |
| HR (95% CI) | Model 1a | | 1 (Reference) | | 1 (Reference) |
| | Model 1b | | 1 (Reference) | | 1 (Reference) |
| ***Ovarian*** | | | | | |
| Cases | | | | 26 | 62 |
| IR | | | | 52 (34-75) | 40 (30-50) |
| HR (95% CI) | Model 1a | | | | 1 (Reference) |
| | Model 1b | | | | 1 (Reference) |

**Supplementary Table 4 Pooled hazard ratios (HRs) and 95% confidence intervals (CIs) for obesity-related cancer (ORC) outcomes in men and women with new-onset type 2 diabetes mellitus (T2DM) after performing multiple imputation for missing data and generating 10 imputed datasets, matching on waist circumference (WC) with a 0.1 standard deviation calliper, on age with an interval <5 years, and stratifying on matched set.** Cox model 1a is adjusted for WC, smoking, alcohol, Townsend deprivation, and Summed Metabolic Equivalent Task (MET) minutes per week for all activity by including them as covariates. Cox model 1b adjusted similarly, but also time-split at 1 year with an interaction term added between T2DM and time, to account for detection-time bias and reverse causality.

| | | **Men** | | **Women** | |
|---|---|---|---|---|---|
| | | **New-onset T2DM** | **Matched controls** | **New-onset T2DM** | **Matched controls** |
| Participants | | 14,094 | 42,205 | 9767 | 29,246 |
| Person-years | | 74,370 | 226,079 | 50,608 | 153,077 |
| ***ORC*** | | | | | |
| Cases | | 561 | 891 | 486 | 1040 |
| IR | | 754 (693-819) | 394 (369-421) | 960 (877-1050) | 679 (639-722) |
| HR (95% CI) | Model 1a | 1.84 (1.65-2.06) | 1 (Reference) | 1.41 (1.26-1.59) | 1 (Reference) |
| | Model 1b | 1.32 (1.15-1.51) | 1 (Reference) | 1.20 (1.05-1.38) | 1 (Reference) |
| ***Liver*** | | | | | |
| Cases | | 71 | 41 | 13 | 5 |
| IR | | 95 (75-120) | 18 (13-25) | 26 (14-44) | 3 (1-8) |
| HR (95% CI) | Model 1a | 5.79 (3.63-9.24) | 1 (Reference) | | 1 (Reference) |
| | Model 1b | 4.03 (2.44-6.64) | 1 (Reference) | | 1 (Reference) |
| ***Pancreas*** | | | | | |
| Cases | | 102 | 90 | 63 | 41 |
| IR | | 137 (112-166) | 40 (32-39) | 124 (96-159) | 27 (19-36) |
| HR (95% CI) | Model 1a | 3.70 (2.70-5.07) | 1 (Reference) | 4.36 (2.80-6.77) | 1 (Reference) |
| | Model 1b | 1.41 (0.93-2.14) | 1 (Reference) | 2.35 (1.36-4.06) | 1 (Reference) |
| ***Colorectum*** | | | | | |
| Cases | | 208 | 421 | 83 | 183 |
| IR | | 280 (243-320) | 186 (169-205) | 164 (131-203) | 120 (103-138) |
| HR (95% CI) | Model 1a | 1.45 (1.21-1.73) | 1 (Reference) | 1.34 (1.02-1.76) | 1 (Reference) |
| | Model 1b | 1.10 (0.89-1.36) | 1 (Reference) | 1.24 (0.90-1.71) | 1 (Reference) |
| ***Postmenopausal breast*** | | | | | |
| Cases | | | | 127 | 412 |
| IR | | | | 251 (209-299) | 269 (244-296) |
| HR (95% CI) | Model 1a | | | 0.97 (0.79-1.20) | 1 (Reference) |
| | Model 1b | | | 1.01 (0.81-1.28) | 1 (Reference) |
| ***Endometrium*** | | | | | |
| Cases | | | | 78 | 154 |
| IR | | | | 154 (122-192) | 101 (85-118) |
| HR (95% CI) | Model 1a | | | 1.70 (1.26-2.31) | 1 (Reference) |
| | Model 1b | | | 1.33 (0.93-1.89) | 1 (Reference) |
| ***Kidney*** | | | | | |
| Cases | | 82 | 122 | 24 | 54 |

| | | | | | |
|---|---|---|---|---|---|
| IR | | 110 (88-137) | 54 (45-64) | 47 (30-71) | 35 (27-46) |
| HR (95% CI) | Model 1a | 1.91 (1.41-2.58) | 1 (Reference) | | 1 (Reference) |
| | Model 1b | 1.50 (1.06-2.14) | 1 (Reference) | | 1 (Reference) |
| ***Oesophagus*** | | | | | |
| Cases | | 33 | 71 | 9 | 10 |
| IR | | 44 (31-62) | 31 (25-40) | 18 (8-34) | 7 (3-12) |
| HR (95% CI) | Model 1a | 1.35 (0.86-2.12) | 1 (Reference) | | 1 (Reference) |
| | Model 1b | 1.01 (0.60-1.70) | 1 (Reference) | | 1 (Reference) |
| ***Gastric cardia*** | | | | | |
| Cases | | 13 | 41 | 5 | 7 |
| IR | | 17 (9-30) | 18 (13-25) | 10 (3-23) | 5 (2-9) |
| HR (95% CI) | Model 1a | | 1 (Reference) | | 1 (Reference) |
| | Model 1b | | 1 (Reference) | | 1 (Reference) |
| ***Multiple myeloma*** | | | | | |
| Cases | | 32 | 68 | 17 | 26 |
| IR | | 43 (29-61) | 30 (23-38) | 34 (20-54) | 17 (11-25) |
| HR (95% CI) | Model 1a | 1.47 (0.92-2.36) | 1 (Reference) | | 1 (Reference) |
| | Model 1b | 1.23 (0.70-2.16) | 1 (Reference) | | 1 (Reference) |
| ***Gallbladder*** | | | | | |
| Cases | | 3 | 9 | 6 | 10 |
| IR | | 4 (1-12) | 4 (2-8) | 12 (4-26) | 7 (3-12) |
| HR (95% CI) | Model 1a | | 1 (Reference) | | 1 (Reference) |
| | Model 1b | | 1 (Reference) | | 1 (Reference) |
| ***Thyroid*** | | | | | |
| Cases | | 6 | 12 | 19 | 26 |
| IR | | 8 (3-17) | 5 (3-9) | 38 (23-59) | 17 (11-25) |
| HR (95% CI) | Model 1a | | 1 (Reference) | | 1 (Reference) |
| | Model 1b | | 1 (Reference) | | 1 (Reference) |
| ***Meningioma*** | | | | | |
| Cases | | 5 | 11 | 16 | 28 |
| IR | | 7 (2-16) | 5 (2-9) | 32 (18-51) | 18 (12-26) |
| HR (95% CI) | Model 1a | | 1 (Reference) | | 1 (Reference) |
| | Model 1b | | 1 (Reference) | | 1 (Reference) |
| ***Ovarian*** | | | | | |
| Cases | | | | 26 | 84 |
| IR | | | | 51 (34-75) | 55 (44-68) |
| HR (95% CI) | Model 1a | | | | 1 (Reference) |
| | Model 1b | | | | 1 (Reference) |

**Supplementary Table 5 Pooled hazard ratios (HRs) and 95% confidence intervals (CIs) for obesity-related cancer (ORC) outcomes in men and women with new-onset type 2 diabetes mellitus (T2DM) after performing multiple imputation for missing data and generating 10 imputed datasets, matching on Waist-to-Hip Ratio (WHR) with a 0.1 standard deviation calliper, on age with an interval <5 years, and stratifying on matched set.** Cox model 1a is adjusted for WHR, smoking, alcohol, Townsend deprivation, and Summed Metabolic Equivalent Task (MET) minutes per week for all activity by including them as covariates. Cox model 1b adjusted similarly, but also time-split at 1 year with an interaction term added between T2DM and time, to account for detection-time bias and reverse causality.

| | | **Men** | | **Women** | |
|---|---|---|---|---|---|
| | | **New-onset T2DM** | **Matched controls** | **New-onset T2DM** | **Matched controls** |
| Participants | | 14,194 | 42,504 | 9786 | 29,328 |
| Person-years | | 74,606 | 227,652 | 50,600 | 154,624 |
| ***ORC*** | | | | | |
| Cases | | 566 | 902 | 487 | 963 |
| IR | | 759 (697-824) | 396 (371-423) | 962 (879-1052) | 623 (584-663) |
| HR (95% CI) | Model 1a | 1.96 (1.75-2.19) | 1 (Reference) | 1.53 (1.37-1.72) | 1 (Reference) |
| | Model 1b | 1.47 (1.28-1.68) | 1 (Reference) | 1.28 (1.11-1.46) | 1 (Reference) |
| ***Liver*** | | | | | |
| Cases | | 72 | 34 | 13 | 3 |
| IR | | 96 (75-121) | 15 (10-21) | 26 (14-44) | 2 (1-6) |
| HR (95% CI) | Model 1a | 6.64 (4.05-10.87) | 1 (Reference) | | 1 (Reference) |
| | Model 1b | 5.48 (3.16-9.49) | 1 (Reference) | | 1 (Reference) |
| ***Pancreas*** | | | | | |
| Cases | | 103 | 83 | 64 | 51 |
| IR | | 138 (113-167) | 36 (29-45) | 126 (97-162) | 33 (25-43) |
| HR (95% CI) | Model 1a | 3.99 (2.93-5.46) | 1 (Reference) | 3.72 (2.48-5.59) | 1 (Reference) |
| | Model 1b | 1.75 (1.16-2.65) | 1 (Reference) | 1.81 (1.09-3.01) | 1 (Reference) |
| ***Colorectum*** | | | | | |
| Cases | | 210 | 427 | 83 | 167 |
| IR | | 281 (245-322) | 188 (170-206) | 164 (131-203) | 108 (92-126) |
| HR (95% CI) | Model 1a | 1.55 (1.29-1.85) | 1 (Reference) | 1.67 (1.26-2.22) | 1 (Reference) |
| | Model 1b | 1.25 (1.01-1.54) | 1 (Reference) | 1.42 (1.02-1.97) | 1 (Reference) |
| ***Postmenopausal breast*** | | | | | |
| Cases | | | | 127 | 404 |
| IR | | | | 251 (209-299) | 261 (236-288) |
| HR (95% CI) | Model 1a | | | 0.99 (0.80-1.22) | 1 (Reference) |
| | Model 1b | | | 1.03 (0.82-1.30) | 1 (Reference) |
| ***Endometrium*** | | | | | |
| Cases | | | | 78 | 111 |
| IR | | | | 154 (122-192) | 72 (59-86) |
| HR (95% CI) | Model 1a | | | 2.03 (1.48-2.79) | 1 (Reference) |
| | Model 1b | | | 1.54 (1.07-2.23) | 1 (Reference) |
| ***Kidney*** | | | | | |
| Cases | | 82 | 134 | 24 | 48 |

| | | | | | |
|---|---|---|---|---|---|
| IR | | 110 (87-136) | 59 (49-69) | 47 (30-71) | 31 (23-41) |
| HR (95% CI) | Model 1a | 1.92 (1.41-2.60) | 1 (Reference) | | 1 (Reference) |
| | Model 1b | 1.63 (1.14-2.33) | 1 (Reference) | | 1 (Reference) |
| ***Oesophagus*** | | | | | |
| Cases | | 33 | 75 | 9 | 13 |
| IR | | 44 (30-62) | 33 (26-41) | 18 (8-34) | 8 (4-14) |
| HR (95% CI) | Model 1a | 1.27 (0.79-2.04) | 1 (Reference) | | 1 (Reference) |
| | Model 1b | 0.97 (0.55-1.71) | 1 (Reference) | | 1 (Reference) |
| ***Gastric cardia*** | | | | | |
| Cases | | 14 | 40 | 5 | 6 |
| IR | | 19 (10-31) | 18 (13-24) | 10 (3-23) | 4 (1-8) |
| HR (95% CI) | Model 1a | | 1 (Reference) | | 1 (Reference) |
| | Model 1b | | 1 (Reference) | | 1 (Reference) |
| ***Multiple myeloma*** | | | | | |
| Cases | | 32 | 64 | 17 | 32 |
| IR | | 43 (29-61) | 28 (22-36) | 34 (20-54) | 21 (14-29) |
| HR (95% CI) | Model 1a | 1.52 (0.95-2.42) | 1 (Reference) | | 1 (Reference) |
| | Model 1b | 1.08 (0.63-1.86) | 1 (Reference) | | 1 (Reference) |
| ***Gallbladder*** | | | | | |
| Cases | | 3 | 5 | 6 | 8 |
| IR | | 4 (1-12) | 2 (1-5) | 12 (4-26) | 5 (2-10) |
| HR (95% CI) | Model 1a | | 1 (Reference) | | 1 (Reference) |
| | Model 1b | | 1 (Reference) | | 1 (Reference) |
| ***Thyroid*** | | | | | |
| Cases | | 6 | 13 | 19 | 17 |
| IR | | 8 (3-17) | 6 (3-10) | 38 (23-59) | 11 (6-18) |
| HR (95% CI) | Model 1a | | 1 (Reference) | | 1 (Reference) |
| | Model 1b | | 1 (Reference) | | 1 (Reference) |
| ***Meningioma*** | | | | | |
| Cases | | 5 | 12 | 16 | 26 |
| IR | | 7 (2-16) | 5 (3-9) | 32 (18-51) | 17 (11-25) |
| HR (95% CI) | Model 1a | | 1 (Reference) | | 1 (Reference) |
| | Model 1b | | 1 (Reference) | | 1 (Reference) |
| ***Ovarian*** | | | | | |
| Cases | | | | 26 | 77 |
| IR | | | | 51 (34-75) | 50 (39-62) |
| HR (95% CI) | Model 1a | | | | 1 (Reference) |
| | Model 1b | | | | 1 (Reference) |

**Supplementary Table 6 Pooled hazard ratios (HRs) and 95% confidence intervals (CIs) for total cancer outcomes in men and women with new-onset type 2 diabetes mellitus (T2DM) after performing multiple imputation for missing data and generating 10 imputed datasets, matching on BMI with a 0.1 standard deviation calliper, on age with an interval <5 years, and stratifying on matched set.** Cox model 1a is adjusted for BMI, smoking, alcohol, Townsend deprivation, and Summed Metabolic Equivalent Task (MET) minutes per week for all activity by including them as covariates. Cox model 1b adjusted similarly, but also time-split at 1 year with an interaction term added between T2DM and time, to account for detection-time bias and reverse causality. Cox model 1b* is adjusted similarly to 1b, but prostate cancer has been excluded from the outcome set.

| | | Men | | Women | |
|---|---|---|---|---|---|
| | | **New-onset T2DM** | **Matched controls** | **New-onset T2DM** | **Matched controls** |
| Participants | | 14,009 | 41,911 | 9762 | 29,259 |
| Person-years | | 73,951 | 224,970 | 50,484 | 153,943 |
| ***Total cancer*** | | | | | |
| Cases (excluding prostate) | | 1590 (1187) | 3553 (2141) | 842 | 1709 |
| IR | | 2150 (2046-2258) | 1579 (1528-1632) | 1668 (1557-1784) | 1110 (1058-1164) |
| IR* | | 1605 (1515-1699) | 952 (912-993) | | |
| HR (95% CI) | Model 1a | 1.35 (1.27-1.44) | 1 (Reference) | 1.50 (1.37-1.64) | 1 (Reference) |
| | Model 1b | 1.06 (0.98-1.14) | 1 (Reference) | 1.32 (1.19-1.46) | 1 (Reference) |
| | Model 1a* | 1.64 (1.51-1.76) | 1 (Reference) | | 1 (Reference) |
| | Model 1b* | 1.21 (1.10-1.32) | 1 (Reference) | | 1 (Reference) |

**Supplementary Table 7 Pooled hazard ratios (HRs) and 95% confidence intervals (CIs) for total cancer outcomes in men and women with new-onset type 2 diabetes mellitus (T2DM) after performing multiple imputation for missing data and generating 10 imputed datasets, matching on waist circumference (WC) with a 0.1 standard deviation calliper, on age with an interval <5 years, and stratifying on matched set.** Cox model 1a is adjusted for WC, smoking, alcohol, Townsend deprivation, and Summed Metabolic Equivalent Task (MET) minutes per week for all activity by including them as covariates. Cox model 1b adjusted similarly, but also time-split at 1 year with an interaction term added between T2DM and time, to account for detection-time bias and reverse causality. Cox model 1b* is adjusted similarly to 1b, but prostate cancer has been excluded from the outcome set.

| | | Men | | Women | |
|---|---|---|---|---|---|
| | | **New-onset T2DM** | **Matched controls** | **New-onset T2DM** | **Matched controls** |
| Participants | | 14,094 | 42,205 | 9767 | 29,246 |
| Person-years | | 74,370 | 226,079 | 50,608 | 153,077 |
| ***Total cancer*** | | | | | |
| Cases (excluding prostate) | | 1598 (1194) | 3595 (2197) | 843 | 1776 |
| IR | | 2149 (2045-2257) | 1590 (1539-1643) | 1666 (1555-1782) | 1160 (1107-1215) |
| IR* | | 1605 (1516-1699) | 972 (932-1013) | | |
| HR (95% CI) | Model 1a | 1.32 (1.24-1.41) | 1 (Reference) | 1.43 (1.31-1.56) | 1 (Reference) |
| | Model 1b | 1.02 (0.95-1.10) | 1 (Reference) | 1.21 (1.09-1.34) | 1 (Reference) |
| | Model 1a* | 1.57 (1.45-1.69) | 1 (Reference) | | 1 (Reference) |
| | Model 1b* | 1.15 (1.05-1.26) | 1 (Reference) | | 1 (Reference) |

**Supplementary Table 8 Pooled hazard ratios (HRs) and 95% confidence intervals (CIs) for total cancer outcomes in men and women with new-onset type 2 diabetes mellitus (T2DM) after performing multiple imputation for missing data and generating 10 imputed datasets, matching on Waist-To-Hip Ratio (WHR) with a 0.1 standard deviation calliper, on age with an interval <5 years, and stratifying on matched set.** Cox model 1a is adjusted for WHR, smoking, alcohol, Townsend deprivation, and Summed Metabolic Equivalent Task (MET) minutes per week for all activity by including them as covariates. Cox model 1b adjusted similarly, but also time-split at 1 year with an interaction term added between T2DM and time, to account for detection-time bias and reverse causality. Cox model 1b* is adjusted similarly to 1b, but prostate cancer has been excluded from the outcome set.

| | | Men | | Women | |
|---|---|---|---|---|---|
| | | **New-onset T2DM** | **Matched controls** | **New-onset T2DM** | **Matched controls** |
| Participants | | 14,194 | 42,504 | 9786 | 29,328 |
| Person-years | | 74,606 | 227,652 | 50,600 | 154,624 |
| ***Total cancer*** | | | | | |
| Cases (excluding prostate) | | 1602 (1199) | 3699 (2207) | 844 | 1720 |
| IR | | 2147 (2043-2555) | 1625 (1573-1678) | 1668 (1557-1784) | 1112 (1060-1166) |
| IR* | | 1607 (1517-1701) | 969 (929-1011) | | |
| HR (95% CI) | Model 1a | 1.33 (1.25-1.42) | 1 (Reference) | 1.50 (1.37-1.64) | 1 (Reference) |
| | Model 1b | 1.03 (0.96-1.11) | 1 (Reference) | 1.24 (1.12-1.38) | 1 (Reference) |
| | Model 1a* | 1.66 (1.54-1.79) | 1 (Reference) | | |
| | Model 1b* | 1.23 (1.12-1.34) | 1 (Reference) | | |

**Supplementary Table 9 Pooled hazard ratios (HRs) and 95% confidence intervals (CIs) for non-obesity-related cancer (NORC) outcomes in men and women with new-onset type 2 diabetes mellitus (T2DM) after performing multiple imputation for missing data and generating 10 imputed datasets, matching on BMI with a 0.1 standard deviation calliper, on age with an interval <5 years, and stratifying on matched set.** Cox model 1a is adjusted for BMI, smoking, alcohol, Townsend deprivation, and Summed Metabolic Equivalent Task (MET) minutes per week for all activity by including them as covariates. Cox model 1b adjusted similarly, but also time-split at 1 year with an interaction term added between T2DM and time, to account for detection-time bias and reverse causality. Cox model 1b* is adjusted similarly to 1b, but prostate cancer has been excluded from the outcome set.

| | | Men | | Women | |
|---|---|---|---|---|---|
| | | **New-onset T2DM** | **Matched controls** | **New-onset T2DM** | **Matched controls** |
| Participants | | 14,009 | 41,911 | 9762 | 29,259 |
| Person-years | | 73,951 | 224,970 | 50,484 | 153,943 |
| ***NORC*** | | | | | |
| Cases (excluding prostate) | | 1029 (626) | 2664 (1252) | 355 | 706 |
| IR | | 1391 (1308-1479) | 1184 (1140-1230) | 703 (632-780) | 459 (425-494) |
| IR* | | 847 (781-915) | 557 (526-588) | | |
| HR (95% CI) | Model 1a | 1.17 (1.08-1.26) | 1 (Reference) | 1.52 (1.33-1.75) | 1 (Reference) |
| | Model 1b | 0.94 (0.86-1.03) | 1 (Reference) | 1.03 (0.88-1.21) | 1 (Reference) |
| | Model 1a* | 1.45 (1.31-1.61) | 1 (Reference) | | |
| | Model 1b* | 1.07 (0.95-1.22) | 1 (Reference) | | |
| ***Lung*** | | | | | |
| Cases | | 157 | 257 | 97 | 134 |
| IR | | 212 (180-248) | 114 (100-128) | 192 (156-234) | 87 (73-103) |
| HR (95% CI) | Model 1a | 1.67 (1.31-2.12) | 1 (Reference) | 2.04 (1.46-2.83) | 1 (Reference) |
| | Model 1b | 1.11 (0.83-1.48) | 1 (Reference) | 1.63 (1.10-2.43) | 1 (Reference) |
| ***Prostate*** | | | | | |
| Cases | | 403 | 1412 | | |
| IR | | 545 (493-601) | 627 (595-660) | | |
| HR (95% CI) | Model 1a | 0.91 (0.81-1.02) | 1 (Reference) | | 1 (Reference) |
| | Model 1b | 0.81 (0.71-0.93) | 1 (Reference) | | 1 (Reference) |
| ***Bladder*** | | | | | |
| Cases | | 66 | 100 | 13 | 19 |
| IR | | 89 (69-114) | 45 (36-53) | 26 (14-44) | 12 (7-19) |
| HR (95% CI) | Model 1a | 1.95 (1.38-2.75) | 1 (Reference) | | 1 (Reference) |
| | Model 1b | 1.36 (0.91-2.04) | 1 (Reference) | | 1 (Reference) |
| ***Melanoma*** | | | | | |
| Cases | | 50 | 181 | 23 | 85 |
| IR | | 68 (50-89) | 81 (69-92) | 46 (29-68) | 55 (44-68) |
| HR (95% CI) | Model 1a | 0.91 (0.65-1.27) | 1 (Reference) | | 1 (Reference) |
| | Model 1b | 0.98 (0.68-1.41) | 1 (Reference) | | 1 (Reference) |
| ***Non-follicular lymphoma*** | | | | | |
| Cases | | 40 | 85 | 12 | 38 |
| IR | | 54 (39-74) | 38 (30-46) | 24 (12-41) | 25 (17-34) |
| HR (95% CI) | Model 1a | 1.43 (0.94-2.18) | 1 (Reference) | | 1 (Reference) |

| | | | | | |
|---|---|---|---|---|---|
| | Model 1b | 1.50 (0.91-2.46) | 1 (Reference) | | 1 (Reference) |

**Supplementary Table 10 Pooled hazard ratios (HRs) and 95% confidence intervals (CIs) for non-obesity-related cancer (NORC) outcomes in men and women with new-onset type 2 diabetes mellitus (T2DM) after performing multiple imputation for missing data and generating 10 imputed datasets, matching on waist circumference (WC) with a 0.1 standard deviation calliper, on age with an interval <5 years, and stratifying on matched set.** Cox model 1a is adjusted for WC, smoking, alcohol, Townsend deprivation, and Summed Metabolic Equivalent Task (MET) minutes per week for all activity by including them as covariates. Cox model 1b adjusted similarly, but also time-split at 1 year with an interaction term added between T2DM and time, to account for detection-time bias and reverse causality. Cox model 1b* is adjusted similarly to 1b, but prostate cancer has been excluded from the outcome set.

| | | **Men** | | **Women** | |
|---|---|---|---|---|---|
| | | **New-onset T2DM** | **Matched controls** | **New-onset T2DM** | **Matched controls** |
| Participants | | 14,094 | 42,205 | 9767 | 29,246 |
| Person-years | | 74,370 | 226,079 | 50,608 | 153,077 |
| ***NORC*** | | | | | |
| Cases (excluding prostate) | | 1037 (633) | 2704 (1306) | 357 | 736 |
| IR | | 1394 (1311-1482) | 1196 (1151-1242) | 705 (634-783) | 481 (447-517) |
| IR* | | 851 (786-920) | 578 (547-610) | | |
| HR (95% CI) | Model 1a | 1.14 (1.06-1.24) | 1 (Reference) | 1.44 (1.26-1.65) | 1 (Reference) |
| | Model 1b | 0.92 (0.84-1.00) | 1 (Reference) | 1.22 (1.04-1.42) | 1 (Reference) |
| | Model 1a* | 1.38 (1.24-1.53) | 1 (Reference) | | |
| | Model 1b* | 1.03 (0.91-1.17) | 1 (Reference) | | |
| ***Lung*** | | | | | |
| Cases | | 157 | 288 | 97 | 150 |
| IR | | 211 (179-247) | 127 (113-143) | 192 (155-234) | 98 (83-115) |
| HR (95% CI) | Model 1a | 1.38 (1.08-1.76) | 1 (Reference) | 1.96 (1.41-2.71) | 1 (Reference) |
| | Model 1b | 0.95 (0.71-1.28) | 1 (Reference) | 1.41 (0.96-2.08) | 1 (Reference) |
| ***Prostate*** | | | | | |
| Cases | | 404 | 1398 | | |
| IR | | 543 (492-599) | 618 (586-652) | | |
| HR (95% CI) | Model 1a | 0.92 (0.82-1.03) | 1 (Reference) | | 1 (Reference) |
| | Model 1b | 0.81 (0.71-0.93) | 1 (Reference) | | 1 (Reference) |
| ***Bladder*** | | | | | |
| Cases | | 66 | 115 | 14 | 24 |
| IR | | 89 (69-113) | 51 (42-61) | 28 (15-46) | 16 (10-23) |
| HR (95% CI) | Model 1a | 1.75 (1.26-2.45) | 1 (Reference) | | 1 (Reference) |
| | Model 1b | 1.46 (0.99-2.16) | 1 (Reference) | | 1 (Reference) |
| ***Melanoma*** | | | | | |
| Cases | | 51 | 181 | 23 | 87 |
| IR | | 69 (51-90) | 80 (69-93) | 45 (29-68) | 57 (46-70) |
| HR (95% CI) | Model 1a | 0.87 (0.62-1.22) | 1 (Reference) | | 1 (Reference) |
| | Model 1b | 0.95 (0.66-1.36) | 1 (Reference) | | 1 (Reference) |

| | | | | | |
|---|---|---|---|---|---|
| ***Non-follicular lymphoma*** | | | | | |
| Cases | | 41 | 103 | 12 | 34 |
| IR | | 55 (40-75) | 46 (37-55) | 24 (12-41) | 22 (15-31) |
| HR (95% CI) | Model 1a | 1.24 (0.83-1.83) | 1 (Reference) | | 1 (Reference) |
| | Model 1b | 1.09 (0.69-1.70) | 1 (Reference) | | 1 (Reference) |

**Supplementary Table 11 Pooled hazard ratios (HRs) and 95% confidence intervals (CIs) for non-obesity-related cancer (NORC) outcomes in men and women with new-onset type 2 diabetes mellitus (T2DM) after performing multiple imputation for missing data and generating 10 imputed datasets, matching on Waist-To-Hip Ratio (WHR) with a 0.1 standard deviation calliper, on age with an interval <5 years, and stratifying on matched set.** Cox model 1a is adjusted for WHR, smoking, alcohol, Townsend deprivation, and Summed Metabolic Equivalent Task (MET) minutes per week for all activity by including them as covariates. Cox model 1b adjusted similarly, but also time-split at 1 year with an interaction term added between T2DM and time, to account for detection-time bias and reverse causality. Cox model 1b* is adjusted similarly to 1b, but prostate cancer has been excluded from the outcome set.

| | | **Men** | | **Women** | |
|---|---|---|---|---|---|
| | | **New-onset T2DM** | **Matched controls** | **New-onset T2DM** | **Matched controls** |
| Participants | | 14,194 | 42,504 | 9786 | 29,328 |
| Person-years | | 74,606 | 227,652 | 50,600 | 154,624 |
| ***NORC*** | | | | | |
| Cases (excluding prostate) | | 1036 (633) | 2797 (1305) | 357 | 757 |
| IR | | 1389 (1305-1476) | 1229 (1184-1275) | 706 (634-783) | 490 (455-526) |
| IR* | | 848 (784-917) | 573 (543-605) | | |
| HR (95% CI) | Model 1a | 1.13 (1.05-1.22) | 1 (Reference) | 1.46 (1.28-1.68) | 1 (Reference) |
| | Model 1b | 0.90 (0.82-0.98) | 1 (Reference) | 1.22 (1.04-1.42) | 1 (Reference) |
| | Model 1a* | 1.47 (1.32-1.63) | 1 (Reference) | | |
| | Model 1b* | 1.07 (0.95-1.22) | 1 (Reference) | | |
| ***Lung*** | | | | | |
| Cases | | 159 | 323 | 97 | 194 |
| IR | | 213 (181-249) | 142 (127-158) | 192 (155-234) | 125 (108-144) |
| HR (95% CI) | Model 1a | 1.44 (1.13-1.83) | 1 (Reference) | 1.66 (1.21-2.28 | 1 (Reference) |
| | Model 1b | 0.95 (0.71-1.27) | 1 (Reference) | 1.23 (0.84-1.80) | 1 (Reference) |
| ***Prostate*** | | | | | |
| Cases | | 403 | 1492 | | |
| IR | | 540 (489-596) | 655 (623-690) | | |
| HR (95% CI) | Model 1a | 0.85 (0.76-0.95) | 1 (Reference) | | 1 (Reference) |
| | Model 1b | 0.75 (0.66-0.86) | 1 (Reference) | | 1 (Reference) |
| ***Bladder*** | | | | | |
| Cases | | 66 | 114 | 14 | 20 |
| IR | | 88 (68-113) | 50 (41-60) | 28 (15-46) | 13 (8-20) |
| HR (95% CI) | Model 1a | 1.84 (1.32-2.56) | 1 (Reference) | | 1 (Reference) |
| | Model 1b | 1.45 (0.98-2.14) | 1 (Reference) | | 1 (Reference) |
| ***Melanoma*** | | | | | |
| Cases | | 51 | 163 | 23 | 81 |
| IR | | 68 (51-90) | 72 (61-83) | 45 (29-68) | 52 (42-65) |
| HR (95% CI) | Model 1a | 1.06 (0.75-1.49) | 1 (Reference) | | 1 (Reference) |

| | | | | | |
|---|---|---|---|---|---|
| | Model 1b | 1.07 (0.74-1.55) | 1 (Reference) | | 1 (Reference) |
| ***Non-follicular lymphoma*** | | | | | |
| Cases | | 41 | 83 | 12 | 37 |
| IR | | 55 (39-75) | 36 (29-45) | 24 (12-41) | 24 (17-33) |
| HR (95% CI) | Model 1a | 1.50 (0.99-2.27) | 1 (Reference) | | 1 (Reference) |
| | Model 1b | 1.30 (0.81-2.10) | 1 (Reference) | | 1 (Reference) |

**Supplementary Table 12 Hazard ratios (HRs) and 95% confidence intervals (CIs) for cancer outcomes in men and women after matching on BMI with a 0.1 standard deviation calliper, on age with an interval <5 years, and stratifying on matched set.** Hazard ratios were produced using a sub distribution hazard model on a randomly selected imputed dataset to account for competing risks and all models were adjusted for BMI, smoking, alcohol, Townsend deprivation, and Summed Metabolic Equivalent Task (MET) minutes per week for all activity by including them as covariates. Incidence rates are reported per 100,000 person-years. All models were left truncated based on follow-up time >1 year to account for detection bias and reverse causality. Model 1* refers to a where prostate cancer was excluded from the outcome set.

| | | Men | | Women | |
|---|---|---|---|---|---|
| | | **New-onset T2DM** | **Matched controls** | **New-onset T2DM** | **Matched controls** |
| Participants | | 14,009 | 41,911 | 9762 | 29,259 |
| Person-years | | 73,951 | 224,970 | 50,484 | 153,943 |
| ***Total cancer*** | | | | | |
| Events | | 1590 (1187) | 3553 (2141) | 842 | 1709 |
| Competing events | | 1722 | 2374 | 815 | 876 |
| IR | | 2150 (2046-2258) | 1579 (1528-1632) | 1668 (1557-1784) | 1110 (1058-1164) |
| HR (95% CI) | Model 1a | 1.30 (1.24-1.37) | 1 (Reference) | 1.45 (1.35-1.56) | 1 (Reference) |
| | Model 1b | 1.03 (0.97-1.10) | 1 (Reference) | 1.29 (1.18-1.41) | 1 (Reference) |
| | Model 1a* | 1.57 (1.48-1.67) | 1 (Reference) | | |
| | Model 1b* | 1.19 (1.10-1.28) | 1 (Reference) | | |
| ***ORC*** | | | | | |
| Events | | 561 | 889 | 487 | 1003 |
| Competing events | | 2751 | 5038 | 1151 | 1604 |
| IR | | 759 (697-824) | 395 (370-422) | 965 (881-1054) | 652 (612-693) |
| HR (95% CI) | Model 1a | 1.79 (1.64-1.96) | 1 (Reference) | 1.42 (1.30-1.56) | 1 (Reference) |
| | Model 1b | 1.37 (1.23-1.54) | 1 (Reference) | 1.29 (1.15-1.44) | 1 (Reference) |
| ***NORC*** | | | | | |
| Events | | 1029 (626) | 2664 (1252) | 355 | 706 |
| Competing events | | 2283 | 3263 | 1170 | 1582 |
| IR | | 1391 (1308-1479) | 1184 (1140-1230) | 703 (632-780) | 459 (425-494) |
| HR (95% CI) | Model 1a | 1.12 (1.05-1.19) | 1 (Reference) | 1.44 (1.28-1.61) | 1 (Reference) |
| | Model 1b | 0.92 (0.85-0.99) | 1 (Reference) | 1.28 (1.12-1.46) | 1 (Reference) |
| | Model 1a* | 1.38 (1.27-1.50) | 1 (Reference) | | |
| | Model 1b* | 1.06 (0.95-1.17) | 1 (Reference) | | |

**Supplementary Table 13 Hazard ratios (HRs) and 95% confidence intervals (CIs) for cancer outcomes in men and women after matching on waist circumference (WC) with a 0.1 standard deviation calliper, on age with an interval <5 years, and stratifying on matched set.** Hazard ratios were produced using a sub distribution hazard model on a randomly selected imputed dataset to account for competing risks and all models were adjusted for WC, smoking, alcohol, Townsend deprivation, and Summed Metabolic Equivalent Task (MET) minutes per week for all activity by including them as covariates. Incidence rates are reported per 100,000 person-years. All models were left truncated based on follow-up time >1 year to account for detection bias and reverse causality. Model 1* refers to a where prostate cancer was excluded from the outcome set.

| | | Men | | Women | |
|---|---|---|---|---|---|
| | | **New-onset T2DM** | **Matched controls** | **New-onset T2DM** | **Matched controls** |
| Participants | | 14,094 | 42,205 | 9767 | 29,246 |
| Person-years | | 74,370 | 226,079 | 50,608 | 153,077 |
| ***Total cancer*** | | | | | |
| Events | | 1598 (1194) | 3595 (2197) | 843 | 1776 |
| Competing events | | 1747 | 2517 | 817 | 1079 |
| IR | | 2149 (2045-2257) | 1590 (1539-1643) | 1666 (1555-1782) | 1160 (1107-1215) |
| HR (95% CI) | Model 1a | 1.29 (1.22-1.35) | 1 (Reference) | 1.40 (1.30-1.50) | 1 (Reference) |
| | Model 1b | 1.01 (0.95-1.08) | 1 (Reference) | 1.20 (1.11-1.31) | 1 (Reference) |
| | Model 1a* | 1.53 (1.44-1.63) | 1 (Reference) | | |
| | Model 1b* | 1.15 (1.07-1.24) | 1 (Reference) | | |
| ***ORC*** | | | | | |
| Events | | 561 | 891 | 486 | 1040 |
| Competing events | | 2784 | 5221 | 1157 | 1755 |
| IR | | 754 (693-819) | 394 (369-421) | 960 (877-1050) | 679 (639-722) |
| | Model 1a | 1.79 (1.64-1.96) | 1 (Reference) | 1.37 (1.25-1.51) | 1 (Reference) |
| HR (95% CI) | Model 1b | 1.31 (1.17-1.46) | 1 (Reference) | 1.20 (1.08-1.35) | 1 (Reference) |
| ***NORC*** | | | | | |
| Events | | 1037 (633) | 2704 (1306) | 357 | 736 |
| Competing events | | 2308 | 3408 | 1303 | 2119 |
| IR | | 1394 (1311-1482) | 1196 (1151-1242) | 705 (634-783) | 481 (447-517) |
| HR (95% CI) | Model 1a | 1.10 (1.03-1.17) | 1 (Reference) | 1.41 (1.26-1.57) | 1 (Reference) |
| | Model 1b | 0.91 (0.84-0.98) | 1 (Reference) | 1.22 (1.07-1.39) | 1 (Reference) |
| | Model 1a* | 1.32 (1.21-1.43) | 1 (Reference) | | |
| | Model 1b* | 1.03 (0.93-1.14) | 1 (Reference) | | |

**Supplementary Table 14 Hazard ratios (HRs) and 95% confidence intervals (CIs) for cancer outcomes in men and women after matching on Waist-To-Hip Ratio (WHR) with a 0.1 standard deviation calliper, on age with an interval <5 years, and stratifying on matched set.** Hazard ratios were produced using a sub distribution hazard model on a randomly selected imputed dataset to account for competing risks and all models were adjusted for WHR, smoking, alcohol, Townsend deprivation, and Summed Metabolic Equivalent Task (MET) minutes per week for all activity by including them as covariates. Incidence rates are reported per 100,000 person-years. All models were left truncated based on follow-up time >1 year to account for detection bias and reverse causality. Model 1* refers to a where prostate cancer was excluded from the outcome set.

| | | Men | | Women | |
|---|---|---|---|---|---|
| | | **New-onset T2DM** | **Matched controls** | **New-onset T2DM** | **Matched controls** |
| Participants | | 14,194 | 42,504 | 9786 | 29,328 |
| Person-years | | 74,606 | 227,652 | 50,600 | 154,624 |
| ***Total cancer*** | | | | | |
| Events (Excluding prostate) | | 1602 (1199) | 3699 (2207) | 844 | 1720 |
| Competing events | | 1758 | 2605 | 816 | 1002 |
| IR | | 2147 (2043-2555) | 1625 (1573-1678) | 1668 (1557-1784) | 1112 (1060-1166) |
| HR (95% CI) | Model 1a | 1.30 (1.23-1.37) | 1 (Reference) | 1.46 (1.36-1.57) | 1 (Reference) |
| | Model 1b | 1.02 (0.96-1.09) | 1 (Reference) | 1.22 (1.12-1.33) | 1 (Reference) |
| | Model 1a* | 1.61 (1.51-1.71) | 1 (Reference) | | |
| | Model 1b* | 1.22 (1.13-1.31) | 1 (Reference) | | |
| ***ORC*** | | | | | |
| Events | | 566 | 902 | 487 | 963 |
| Competing events | | 2794 | 5402 | 1156 | 1687 |
| IR | | 759 (697-824) | 396 (371-423) | 962 (879-1052) | 623 (584-663) |
| | Model 1a | 1.88 (1.71-2.06) | 1 (Reference) | 1.48 (1.34-1.63) | 1 (Reference) |
| HR (95% CI) | Model 1b | 1.46 (1.30-1.63) | 1 (Reference) | 1.25 (1.12-1.40) | 1 (Reference) |
| ***NORC*** | | | | | |
| Events | | 1036 (633) | 2797 (1305) | 357 | 757 |
| Competing events | | 2324 | 3507 | 1173 | 1556 |
| IR | | 1389 (1305-1476) | 1229 (1184-1275) | 706 (634-783) | 490 (455-526) |
| HR (95% CI) | Model 1a | 1.09 (1.02-1.16) | 1 (Reference) | 1.42 (1.26-1.59) | 1 (Reference) |
| | Model 1b | 0.89 (0.82-0.96) | 1 (Reference) | 1.18 (1.03-1.35) | 1 (Reference) |
| | Model 1a* | 1.37 (1.26-1.49) | 1 (Reference) | | |
| | Model 1b* | 1.05 (0.94-1.16) | 1 (Reference) | | |

**Supplementary Table 15 Top 10 causes of mortality in men with new-onset T2DM during the period from recruitment (2006-2010) to the latest linkage of the death registry (31 May 2024 for England and Wales, and 31 December 2023 for Scotland). N=14,186**. All causes of mortality are reported using the 10th revision of the International Classification of Diseases (ICD-10) coding.

| Cause of mortality | Men with new-onset T2DM | | |
|---|---|---|---|
| | Number of events | Percentage of all deaths (%) | Cause-specific mortality (per 100,000 person years) |
| **1**. I21.9 Acute myocardial infarction, unspecified | 174 | 6.87 | 235.29 |
| **2.** I25.9 Chronic ischaemic heart disease, unspecified | 170 | 6.71 | 229.88 |
| **3.** U07.1 COVID-19 virus identified | 170 | 6.71 | 229.88 |
| **4.** C34.9 Bronchus or lung, unspecified | 126 | 4.97 | 170.38 |
| **5.** I25.1 Atherosclerotic heart disease | 115 | 4.54 | 155.51 |
| **6.** C25.9 Pancreas, unspecified | 105 | 4.14 | 141.99 |
| **7.** J84.1 Other interstitial pulmonary diseases with fibrosis | 63 | 2.49 | 85.19 |
| **8.** C22.0 Liver cell carcinoma | 57 | 2.25 | 77.08 |
| **9.** J44.0 Chronic obstructive pulmonary disease with acute lower respiratory infection | 50 | 1.97 | 67.61 |
| **10.** I64 Stroke, not specified as haemorrhage or infarction | 49 | 1.93 | 66.26 |

**Supplementary Table 16 Top 10 causes of mortality in unexposed men during the time from recruitment (2006-2010) to the latest linkage of the death registry (31 May 2024 for England and Wales, and 31 December 2023 for Scotland). N=190,506**. All causes of mortality are reported using the 10th revision of the International Classification of Diseases (ICD-10) coding.

| Cause of mortality | Unexposed men | | |
|---|---|---|---|
| | Number of events | Percentage of all deaths (%) | Cause-specific mortality (per 100,000 person years) |
| **1.** I21.9 Acute myocardial infarction, unspecified | 234 | 6.51 | 104.01 |
| **2.** C34.9 Bronchus or lung, unspecified | 223 | 6.21 | 99.12 |
| **3.** U07.1 COVID-19 virus identified | 211 | 5.87 | 93.79 |
| **4.** I25.9 Chronic ischaemic heart disease, unspecified | 181 | 5.04 | 80.46 |
| **5.** I25.1 Atherosclerotic heart disease | 164 | 4.56 | 72.90 |
| **6.** C15.9 Oesophagus, unspecified | 103 | 2.87 | 45.78 |
| **7.** C25.9 Pancreas, unspecified | 103 | 2.87 | 45.78 |
| **8.** C61 Malignant neoplasm of prostate | 97 | 2.70 | 43.12 |
| **9.** J84.1 Other interstitial pulmonary diseases with fibrosis | 81 | 2.25 | 36.00 |
| **10.** F03 Unspecified dementia | 77 | 2.14 | 34.23 |

**Supplementary Table 17 Top 10 causes of mortality in women with new-onset T2DM during the period from recruitment (2006-2010) to the latest linkage of the death registry (31 May 2024 for England and Wales, and 31 December 2023 for Scotland). N=9775**. All causes of mortality are reported using the 10th revision of the International Classification of Diseases (ICD-10) coding.

| Cause of mortality | Women with new-onset T2DM | | |
|---|---|---|---|
| | Number of events | Percentage of all deaths (%) | Cause-specific mortality (per 100,000 person years) |
| U07.1 COVID-19 virus identified | 76 | 6.39 | 150.54 |
| C34.9 Bronchus or lung, unspecified | 75 | 6.31 | 148.56 |
| C25.9 Pancreas, unspecified | 67 | 5.63 | 132.72 |
| I21.9 Acute myocardial infarction, unspecified | 52 | 4.37 | 103.00 |
| I25.9 Chronic ischaemic heart disease, unspecified | 45 | 3.78 | 89.14 |
| I64 Stroke, not specified as haemorrhage or infarction | 33 | 2.78 | 65.37 |
| I25.1 Atherosclerotic heart disease | 29 | 2.44 | 57.44 |
| F03 Unspecified dementia | 25 | 2.10 | 49.52 |
| G30.9 Alzheimer's disease, unspecified | 25 | 2.10 | 49.52 |
| C50.9 Breast, unspecified | 24 | 2.02 | 47.54 |

**Supplementary Table 18 Top 10 causes of mortality in unexposed women during the time from recruitment (2006-2010) to the latest linkage of the death registry (31 May 2024 for England and Wales, and 31 December 2023 for Scotland). N=232,298.** All causes of mortality are reported using the 10th revision of the International Classification of Diseases (ICD-10) coding.

| Cause of mortality | Unexposed women | | |
|---|---|---|---|
| | Number of events | Percentage of all deaths (%) | Cause-specific mortality (per 100,000 person years) |
| C34.9 Bronchus or lung, unspecified | 100 | 6.60 | 64.96 |
| U07.1 COVID-19 virus identified | 89 | 5.87 | 57.81 |
| C25.9 Pancreas, unspecified | 73 | 4.82 | 47.42 |
| I21.9 Acute myocardial infarction, unspecified | 51 | 3.37 | 33.13 |
| C50.9 Breast, unspecified | 38 | 2.51 | 24.68 |
| G30.9 Alzheimer's disease, unspecified | 37 | 2.44 | 24.03 |
| I25.9 Chronic ischaemic heart disease, unspecified | 34 | 2.24 | 22.09 |
| C56 Malignant neoplasm of ovary | 32 | 2.11 | 20.79 |
| C18.9 Colon, unspecified | 25 | 1.65 | 16.24 |
| I25.1 Atherosclerotic heart disease | 25 | 1.65 | 16.24 |

**Supplementary Table 19 Chi-squared statistics and P values after testing the proportional hazards assumption for type 2 diabetes mellitus (T2DM) using the Grambsch and Therneau method based on Schoenfeld residuals in a randomly selected imputed dataset (non-timesplit).** Chi-squared and P values reported are reported for the T2DM covariate in our main Cox models that were time-split, with prostate excluded from total cancer and non-obesity-related cancer outcomes. P values < 0.05 indicates violation of the assumption.

| Cancer | Chi-squared statistic | P values |
|---|---|---|
| Total (men) | 75.14 | $2×10^{-16}$ |
| Total (women) | 28.14 | $1×10^{-7}$ |
| Obesity-related (men) | 22.59 | $2×10^{-6}$ |
| Obesity-related (women) | 11.18 | $8×10^{-4}$ |
| Non-obesity-related(men) | 37.50 | $9×10^{-10}$ |
| Non-obesity-related (women) | 16.31 | $5×10^{-4}$ |

**Supplementary Table 20 Chi-squared statistics and P values after testing the proportional hazards assumption for type 2 diabetes mellitus (T2DM) using the Grambsch and Therneau method based on Schoenfeld residuals in a randomly selected imputed dataset (timesplit).** Chi-squared and P values are reported for the T2DM covariate in our main Cox models that were time-split, with prostate excluded from total cancer and non-obesity-related cancer outcomes. P values <0.05 indicates violation of the assumption.

| Cancer | Chi-squared statistic | P values |
|---|---|---|
| Total (men) | 0.193 | 0.66 |
| Total (women) | 2.43 | 0.12 |
| Obesity-related (men) | 0.0316 | 0.86 |
| Obesity-related (women) | 0.457 | 0.50 |
| Non-obesity-related (men) | 0.502 | 0.48 |
| Non-obesity-related (women) | 2.30 | 0.13 |

**Supplementary Table 21 Incidence rates (IRs) and crude incidence rate ratio (IRR) estimates for total cancer within each yearly interval of follow-up.** Follow-up was divided into one-year intervals and IRs were estimated by dividing the number of events that occurred within the interval by the number of person years contributed to that interval. IRs are reported per 100,000 person years. We estimated 95% confidence intervals (CIs) for IRs assuming a Poisson distribution, using exact methods based on the Chi-squared distribution. Crude IRRs were calculated as the ratio of IRs between participants with and without new-onset type 2 diabetes mellitus (T2DM), and 95% CIs were estimated using a normal approximation on the logarithmic scale.

| | | Men | | Women | |
|---|---|---|---|---|---|
| | | **New-onset T2DM** | **Matched controls** | **New-onset T2DM** | **Matched controls** |
| Participants | | 14,009 | 41,911 | 9762 | 29,259 |
| Person-years | | 73,951 | 224,970 | 50,484 | 153,943 |
| ***Total cancer*** | | | | | |
| 0-1 years | Number of cases | 435 | 359 | 248 | 335 |
| | IR | 3348 | 907 | 2718 | 1220 |
| | IRR | 3.69 (3.21-4.24) | 1 (Reference) | 2.23 (1.89-2.63) | 1 (Reference) |
| 1-2 years | Number of cases | 126 | 325 | 111 | 256 |
| | IR | 1081 | 912 | 1362 | 1035 |
| | IRR | 1.18 (0.96-1.45) | 1 (Reference) | 1.32 (1.05-1.64) | 1 (Reference) |
| 2-3 years | Number of cases | 110 | 305 | 103 | 221 |
| | IR | 1068 | 971 | 1439 | 1015 |
| | IRR | 1.10 (0.88-1.37) | 1 (Reference) | 1.42 (1.12-1.79) | 1 (Reference) |
| 3-4 years | Number of cases | 108 | 269 | 93 | 208 |
| | IR | 1203 | 983 | 1499 | 1098 |
| | IRR | 1.22 (0.98-1.53) | 1 (Reference) | 1.37 (1.07-1.74) | 1 (Reference) |
| 4-5 years | Number of cases | 93 | 222 | 74 | 181 |
| | IR | 1213 | 953 | 1416 | 1127 |
| | IRR | 1.27 (1.00-1.62) | 1 (Reference) | 1.26 (0.96-1.65) | 1 (Reference) |
| 5-6 years | Number of cases | 86 | 195 | 62 | 156 |
| | IR | 1343 | 1007 | 1444 | 1186 |
| | IRR | 1.33 (1.03-1.72) | 1 (Reference) | 1.22 (0.91-1.63) | 1 (Reference) |
| 6-7 years | Number of cases | 84 | 137 | 50 | 112 |
| | IR | 1630 | 879 | 1464 | 1076 |
| | IRR | 1.85 (1.41-2.43) | 1 (Reference) | 1.36 (0.97-1.90) | 1 (Reference) |
| 7-8 years | Number of cases | 49 | 116 | 46 | 86 |
| | IR | 1218 | 956 | 1769 | 1082 |
| | IRR | 1.27 (0.91-1.78) | 1 (Reference) | `1.64 (1.14-2.34) | 1 (Reference) |
| 8-9 years | Number of cases | 37 | 77 | 19 | 58 |
| | IR | 1225 | 849 | 996 | 992 |
| | IRR | 1.44 (0.97-2.13) | 1 (Reference) | 1.00 (0.60-1.69) | 1 (Reference) |
| 9-10 years | Number of cases | 30 | 64 | 19 | 59 |
| | IR | 1495 | 1046 | 1483 | 1484 |
| | IRR | 1.43 (0.93-2.21) | 1 (Reference) | 0.99 (0.60-1.68) | 1 (Reference) |

**Supplementary Table 22 Incidence rates (IRs) and crude incidence rate ratio (IRR) estimates for obesity-related cancer within each yearly interval of follow-up.** Follow-up was divided into one-year intervals and IRs were estimated by dividing the number of events that occurred within the interval by the number of person years contributed to that interval. IRs are reported per 100,000 person years. We estimated 95% confidence intervals (CIs) for IRs assuming a Poisson distribution, using exact methods based on the Chi-squared distribution. Crude IRRs were calculated as the ratio of IRs between participants with and without new-onset type 2 diabetes mellitus (T2DM), and 95% CIs were estimated using a normal approximation on the logarithmic scale.

| | | **Men** | | **Women** | |
|---|---|---|---|---|---|
| | | **New-onset T2DM** | **Matched controls** | **New-onset T2DM** | **Matched controls** |
| Participants | | 14,009 | 41,911 | 9762 | 29,259 |
| Person-years | | 73,951 | 224,970 | 50,484 | 153,943 |
| ***Obesity-related cancer*** | | | | | |
| 0-1 years | Number of cases | 205 | 146 | 143 | 210 |
| | IR | 1578 | 369 | 1567 | 765 |
| | IRR | 4.28 (3.46-5.29) | 1 (Reference) | 2.05 (1.66-2.53) | 1 (Reference) |
| 1-2 years | Number of cases | 53 | 134 | 74 | 149 |
| | IR | 455 | 376 | 908 | 603 |
| | IRR | 1.21 (0.88-1.66) | 1 (Reference) | 1.51 (1.14-2.00) | 1 (Reference) |
| 2-3 years | Number of cases | 48 | 135 | 50 | 134 |
| | IR | 466 | 430 | 698 | 615 |
| | IRR | 1.08 (0.78-1.51) | 1 (Reference) | 1.13 (0.82-1.57) | 1 (Reference) |
| 3-4 years | Number of cases | 53 | 104 | 41 | 118 |
| | IR | 590 | 380 | 661 | 623 |
| | IRR | 1.55 (1.12-2.16) | 1 (Reference) | 1.06 (0.74-1.51) | 1 (Reference) |
| 4-5 years | Number of cases | 44 | 89 | 46 | 106 |
| | IR | 574 | 382 | 880 | 660 |
| | IRR | 1.50 (1.05-2.16) | 1 (Reference) | 1.33 (0.94-1.88) | 1 (Reference) |
| 5-6 years | Number of cases | 43 | 81 | 40 | 87 |
| | IR | 672 | 418 | 932 | 661 |
| | IRR | 1.60 (1.11-2.32) | 1 (Reference) | 1.41 (0.97-2.05) | 1 (Reference) |
| 6-7 years | Number of cases | 42 | 54 | 31 | 60 |
| | IR | 815 | 347 | 907 | 576 |
| | IRR | 2.35 (1.57-3.52) | 1 (Reference) | 1.57 (1.02-2.43) | 1 (Reference) |
| 7-8 years | Number of cases | 28 | 47 | 29 | 50 |
| | IR | 696 | 387 | 1115 | 629 |
| | IRR | 1.80 (1.12-2.87) | 1 (Reference) | 1.77 (1.12-2.80) | 1 (Reference) |
| 8-9 years | Number of cases | 22 | 37 | 9 | 36 |
| | IR | 728 | 408 | 472 | 615 |
| | IRR | 1.78 (1.05-3.02) | 1 (Reference) | 0.77 (0.37-1.59) | 1 (Reference) |
| 9-10 years | Number of cases | 14 | 30 | 11 | 31 |
| | IR | 698 | 490 | 859 | 780 |
| | IRR | 1.42 (0.75-2.68) | 1 (Reference) | 1.10 (0.55-2.19) | 1 (Reference) |

**Supplementary Table 23 Incidence rates (IRs) and crude incidence rate ratio (IRR) estimates for non-obesity-related cancer within each yearly interval of follow-up.** Follow-up was divided into one-year intervals and IRs were estimated by dividing the number of events that occurred within the interval by the number of person year contributed in that interval. IRs are reported per 100,000 person years. We estimated 95% confidence intervals (CIs) for IRs assuming a Poisson distribution, using exact methods based on the Chi-squared distribution. Crude IRRs were calculated as the ratio of IRs between participants with and without new-onset type 2 diabetes mellitus (T2DM), and 95% CIs were estimated using a normal approximation on the logarithmic scale.

| | | Men | | Women | |
|---|---|---|---|---|---|
| | | **New-onset T2DM** | **Matched controls** | **New-onset T2DM** | **Matched controls** |
| Participants | | 14,009 | 41,911 | 9762 | 29,259 |
| Person-years | | 73,951 | 224,970 | 50,484 | 153,943 |
| ***Non-obesity-related cancer*** | | | | | |
| 0-1 years | Number of cases | 230 | 213 | 105 | 125 |
| | IR | 1770 | 538 | 1151 | 455 |
| | IRR | 3.29 (2.73-3.96) | 1 (Reference) | 2.53 (1.95-3.28) | 1 (Reference) |
| 1-2 years | Number of cases | 73 | 191 | 37 | 107 |
| | IR | 626 | 536 | 454 | 433 |
| | IRR | 1.17 (0.89-1.53) | 1 (Reference) | 1.05 (0.72-1.52) | 1 (Reference) |
| 2-3 years | Number of cases | 62 | 170 | 53 | 87 |
| | IR | 602 | 541 | 740 | 400 |
| | IRR | 1.11 (0.83-1.49) | 1 (Reference) | 1.85 (1.32-2.61) | 1 (Reference) |
| 3-4 years | Number of cases | 55 | 165 | 52 | 90 |
| | IR | 613 | 603 | 838 | 475 |
| | IRR | 1.02 (0.75-1.38) | 1 (Reference) | 1.76 (1.25-2.48) | 1 (Reference) |
| 4-5 years | Number of cases | 49 | 133 | 28 | 75 |
| | IR | 639 | 571 | 536 | 467 |
| | IRR | 1.12 (0.81-1.55) | 1 (Reference) | 1.15 (0.74-1.77) | 1 (Reference) |
| 5-6 years | Number of cases | 43 | 114 | 22 | 69 |
| | IR | 672 | 589 | 513 | 524 |
| | IRR | 1.14 (0.80-1.62) | 1 (Reference) | 0.98 (0.60-1.58) | 1 (Reference) |
| 6-7 years | Number of cases | 42 | 83 | 19 | 52 |
| | IR | 815 | 533 | 556 | 500 |
| | IRR | 1.53 (1.06-2.21) | 1 (Reference) | 1.13 (0.66-1.88) | 1 (Reference) |
| 7-8 years | Number of cases | 21 | 69 | 17 | 36 |
| | IR | 568 | 522 | 654 | 453 |
| | IRR | 0.92 (0.56-1.50) | 1 (Reference) | 1.44 (0.81-2.57) | 1 (Reference) |
| 8-9 years | Number of cases | 15 | 40 | 10 | 22 |
| | IR | 497 | 441 | 524 | 376 |
| | IRR | 1.13 (0.62-2.04) | 1 (Reference) | 1.39 (0.66-2.94) | 1 (Reference) |
| 9-10 years | Number of cases | 16 | 34 | 8 | 28 |
| | IR | 797 | 556 | 625 | 704 |
| | IRR | 1.44 (0.79-2.60) | 1 (Reference) | 0.89 (0.40-1.95) | 1 (Reference) |

**Supplementary Table 24. Triangulation of evidence for the causal effect of type 2 diabetes mellitus and site-specific cancer risk**

| Cancer type | Published evidence | | | | | Matched cohort analyses | | | Overall interpretation |
|---|---|---|---|---|---|---|---|---|---|
| | Tsilidis et al. (2015) Observational | Ling et al. (2020) Observational | Pearson-Stuttard et al. (2021) Observational | Pearson-Stuttard et al. (2021) MR | Wu et al. (2026) MR | BMI-matched Men/Women | WC-matched Men/Women | WHR-matched Men/Women | |
| **Pancreas** | NS | S | S | S | S | S/NS | NS/S | S/S | **Likely causal** |
| **Liver** | NS | S | S | NE | NS | S/NE | S/NE | S/NE | **Likely causal** |
| **Kidney** | NS | S | S | S | NS | S/NE | S/NE | S/NE | **Likely causal** |
| **Colorectal** | S | S | S | NS | NS | NS/NS | NS/NS | S/S | **Insufficient evidence for causality** |
| **Endometrial** | S | S | S | S | S | NS | NS | S | **Insufficient evidence for causality** |
| **Breast** | S | S | S | S | NS | NS | NS | NS | **Insufficient evidence for causality** |

**Abbreviations:** S, statistically significant association; NS, not statistically significant; NE, not estimated; MR, Mendelian randomization; BMI, body mass index; WC, waist circumference; WHR, waist-to-hip ratio. For paired cohort results, values are presented as men/women.

**References: Tsilidis et al. (2015) (1), Ling et al. (2020) (2), Pearson Stuttard et al. (2021) (3), Wu et al. (2026) (4).**

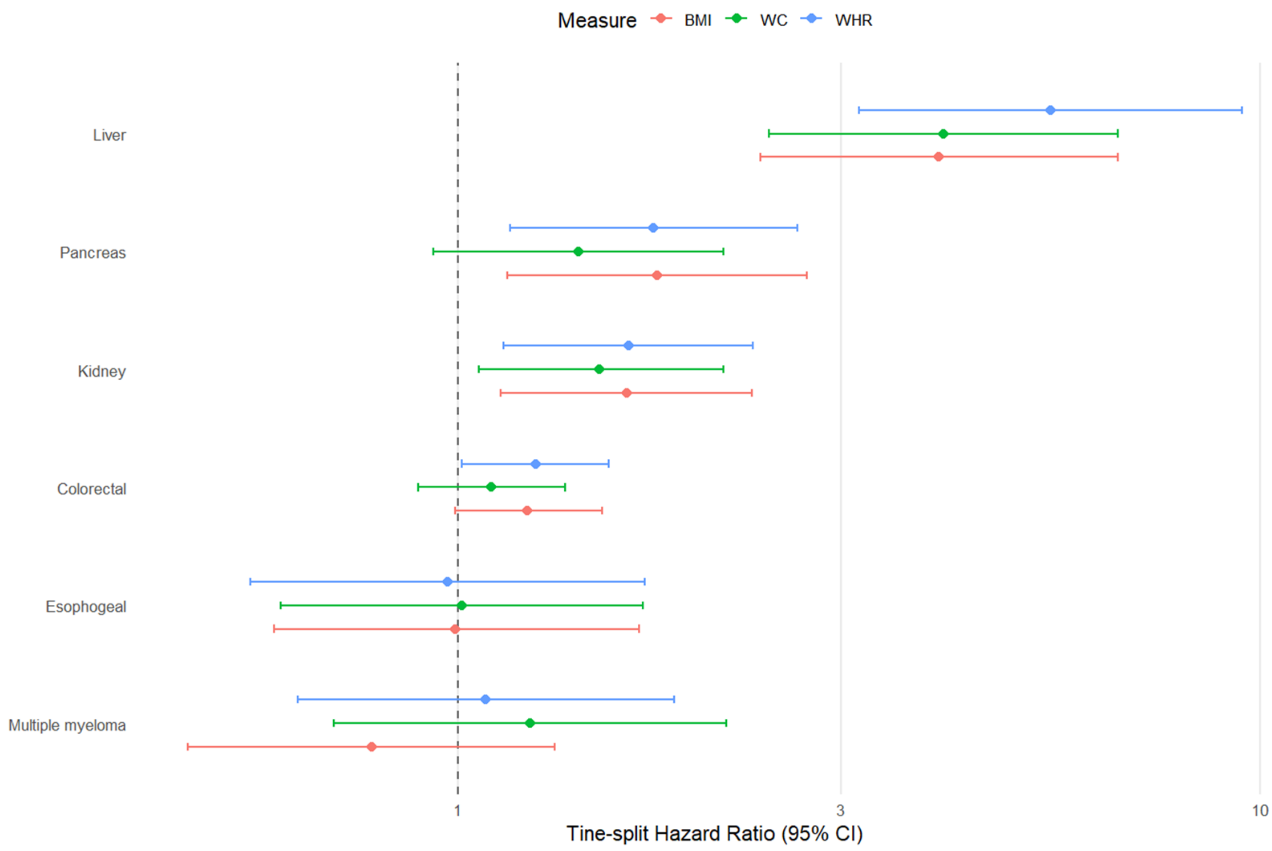

**Supplementary Figure 1 Coefficient plot of time-split hazard ratios (tsHRs) with 95% confidence intervals for the association between new-onset type 2 diabetes and obesity-related cancer outcomes in men, estimated from Cox proportional hazards models using a sex-, age-, and anthropometric measure-matched cohort.** Results are shown for three models: BMI-adjusted, waist circumference (WC), and waist-to-hip ratio (WHR).

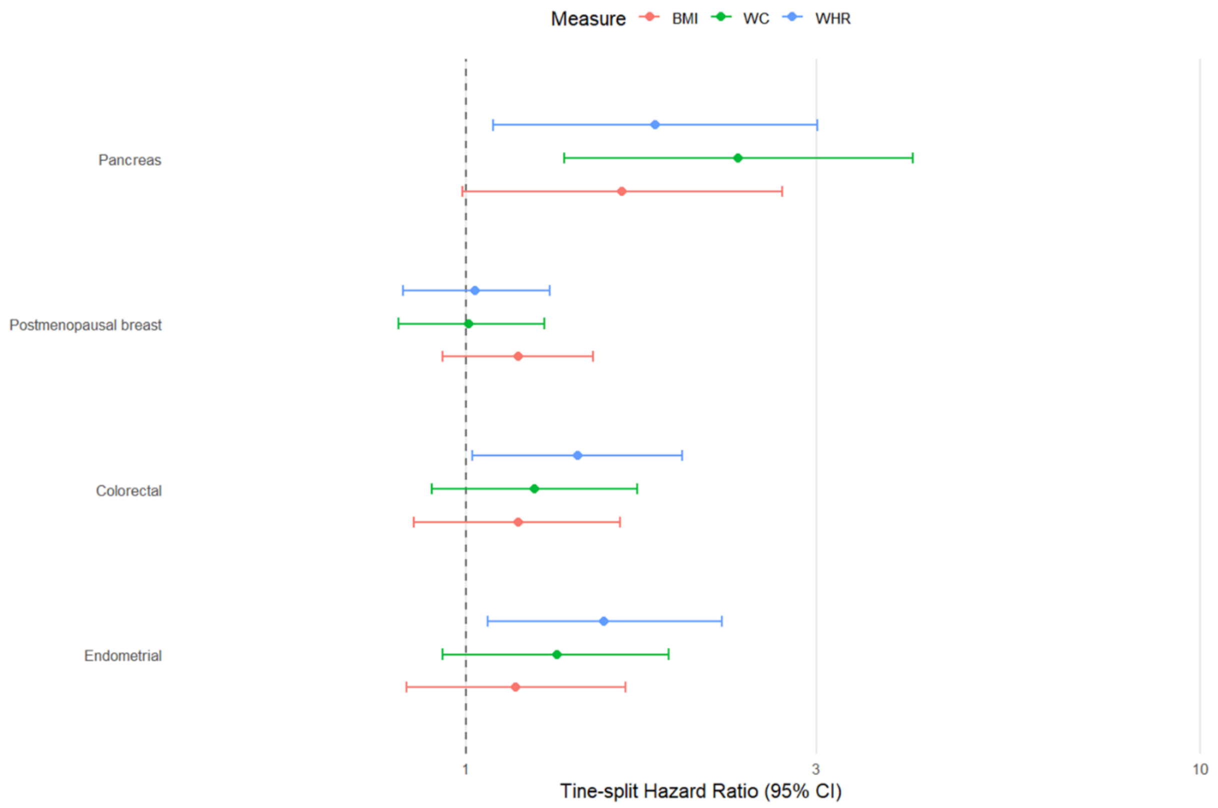

**Supplementary Figure 2 Coefficient plot of time-split hazard ratios (tsHRs) with 95% confidence intervals for the association between new-onset type 2 diabetes and obesity-related cancer outcomes in women, estimated from Cox proportional hazards models using a sex-, age-, and anthropometric measure-matched cohort.** Results are shown for three models: BMI-adjusted, waist circumference (WC), and waist-to-hip ratio (WHR).

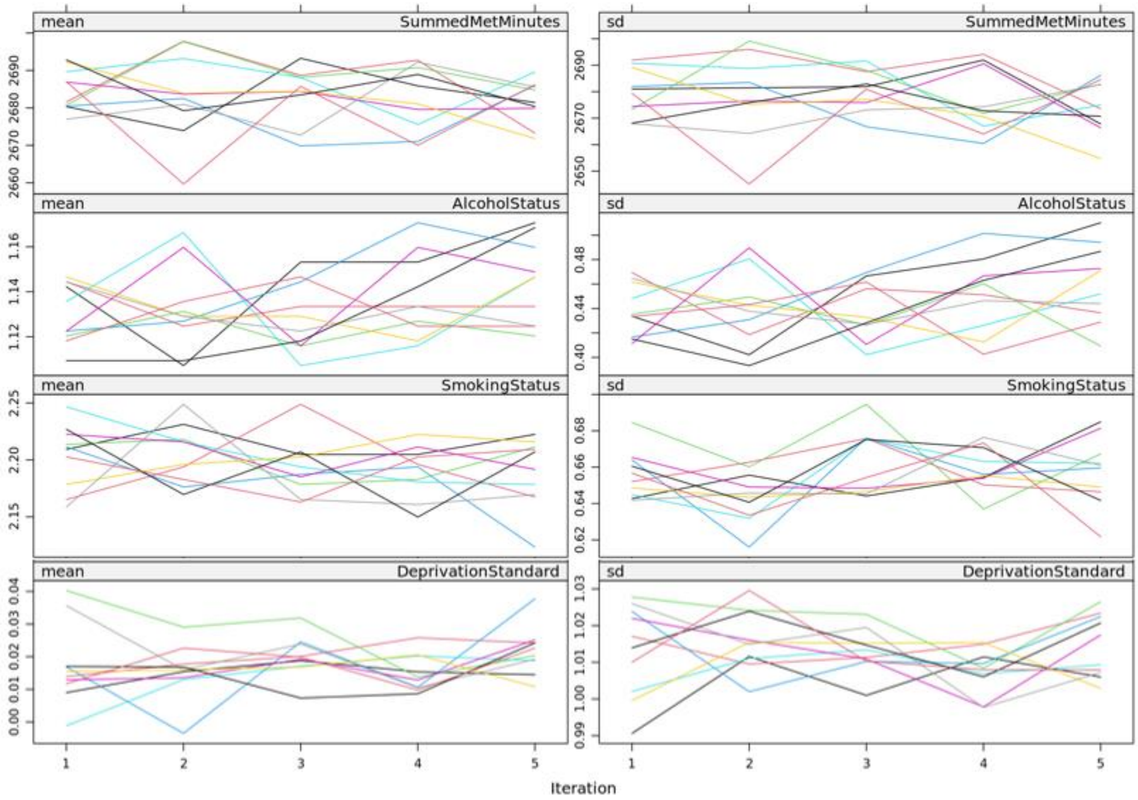

**Supplementary Figure** Error! No text of specified style in document. **Trace plots of the mean (left) and standard deviation (right) of imputed values across five iterations for each imputed dataset.** Each coloured line represents one of the ten imputed datasets.

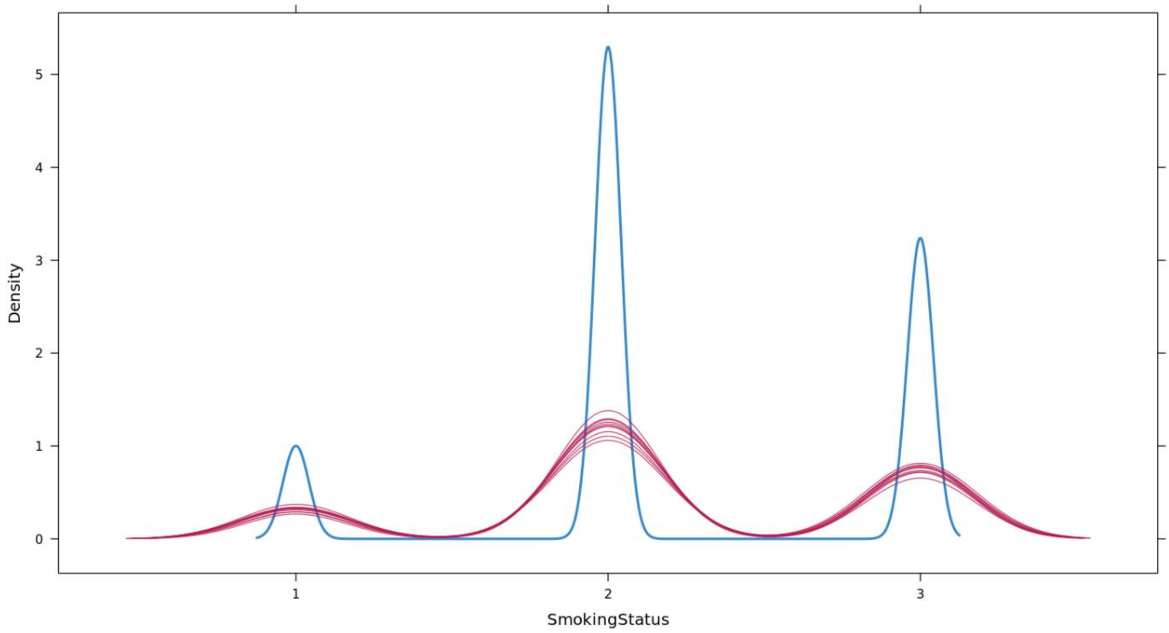

**Supplementary Figure 4 Density plot of smoking status across multiple imputations.** The blue line represents the observed values, and each red line represents imputed values for one of the 10 imputed datasets.

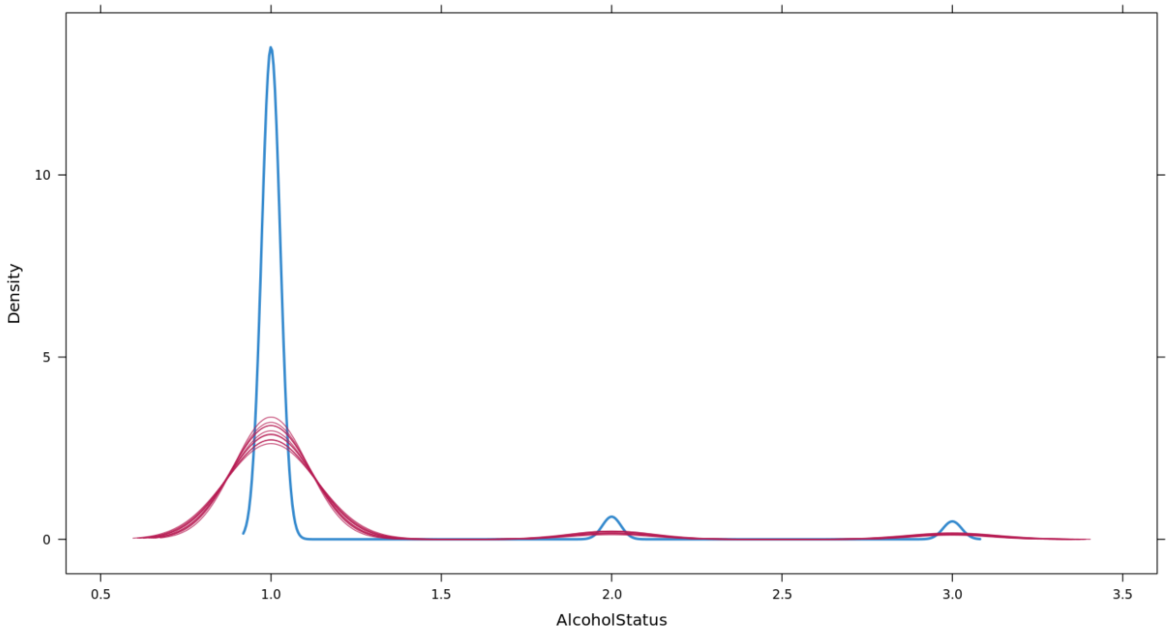

**Supplementary Figure 5 Density plot of Alcohol status across multiple imputations.** The blue line represents the observed values, and each red line represents imputed values for one of the 10 imputed datasets.

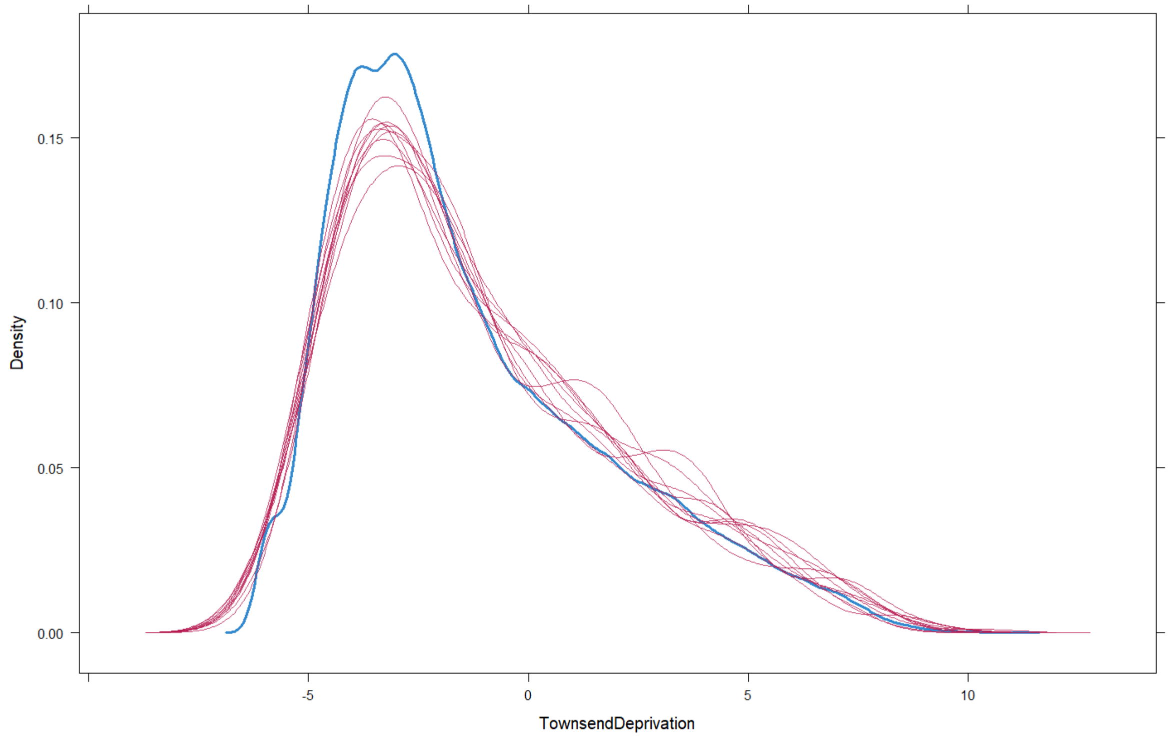

**Supplementary Figure 6 Density plot Townsend deprivation across multiple imputations.** The blue line represents the observed values, and each red line represents imputed values for that imputed dataset.

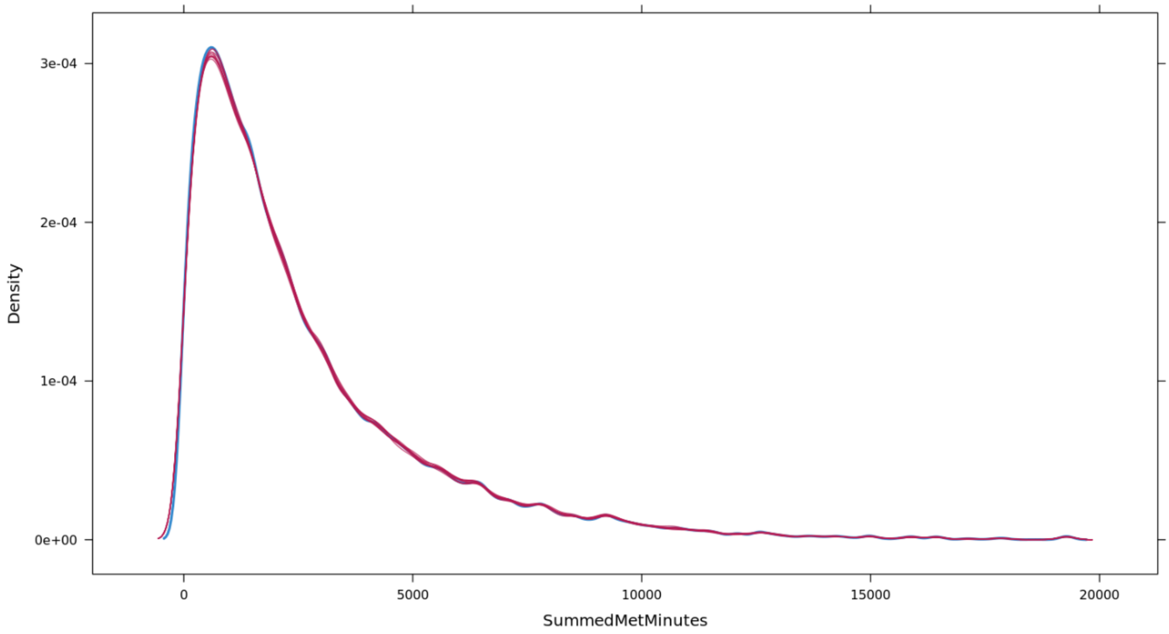

**Supplementary Figure 7 Density plot of Summed Metabolic Equivalent Task (MET) minutes per week for all activity across multiple imputations.** The blue line represents the observed values, and each red line represents imputed values for that imputed dataset.

**References**

1. Tsilidis KK, Kasimis JC, Lopez DS, Ntzani EE, Ioannidis JPA. Type 2 diabetes and cancer: umbrella review of meta-analyses of observational studies. BMJ. 2015;350(jan02 1):g7607-g.

2. Ling S, Brown K, Miksza JK, Howells L, Morrison A, Issa E, et al. Association of type 2 diabetes with cancer: a meta-analysis with bias analysis for unmeasured confounding in 151 cohorts comprising 32 million people. Diabetes care. 2020;43(9):2313-22.

3. Pearson-Stuttard J, Papadimitriou N, Markozannes G, Cividini S, Kakourou A, Gill D, et al. Type 2 Diabetes and Cancer: An Umbrella Review of Observational and Mendelian Randomization Studies. Cancer Epidemiol Biomarkers Prev. 2021;30(6):1218-28.

4. Wu W, Huang G-I, Cui J. Type 2 diabetes mellitus and cancer: A systematic review and meta-analysis of Mendelian randomization studies. Frontiers in Endocrinology. 2026;Volume 17 - 2026.
